# Genome-wide DNA methylation markers associated with metabolic liver cancer

**DOI:** 10.1101/2024.11.15.24317378

**Authors:** Samuel O. Antwi, Ampem Darko Jnr. Siaw, Sebastian M. Armasu, Jacob A. Frank, Irene K. Yan, Fowsiyo Y. Ahmed, Laura Izquierdo-Sanchez, Loreto Boix, Angela Rojas, Jesus M. Banales, Maria Reig, Per Stål, Manuel Romero Gómez, Kirk J. Wangensteen, Amit G. Singal, Lewis R. Roberts, Tushar Patel

## Abstract

**Background and Aims:** Metabolic liver disease is the fastest rising cause of hepatocellular carcinoma (HCC) worldwide, but the underlying molecular processes that drive HCC development in the setting of metabolic perturbations are unclear. We investigated the role of aberrant DNA methylation in metabolic HCC development in a multicenter international study.

**Methods:** We used a case-control design, frequency-matched on age, sex, and study site. Genome-wide profiling of peripheral blood leukocyte DNA was performed using the 850k EPIC array. Cell type proportions were estimated from the methylation data. The study samples were split 80% and 20% for training and validation. Differential methylation analysis was performed with adjustment for cell type, and we generated area under the receiver-operating curves (ROC-AUC).

**Results:** We enrolled 272 metabolic HCC patients and 316 control patients with metabolic liver disease from six sites. Fifty-five differentially methylated CpGs were identified; 33 hypermethylated and 22 hypomethylated in cases versus controls. The panel of 55 CpGs discriminated between cases and controls with AUC=0.79 (95%CI=0.71-0.87), sensitivity=0.77 (95%CI=0.66-0.89), and specificity=0.74 (95%CI=0.64-0.85). The 55-CpG classifier panel performed better than a base model that comprised age, sex, race, and diabetes mellitus (AUC=0.65, 95%CI=0.55-0.75, sensitivity=0.62 (95%CI=0.49-0.75) and specificity=0.64 (95%CI=0.52-0.75). A multifactorial model that combined the 55 CpGs with age, sex, race, and diabetes, yielded AUC=0.78 (95%CI=0.70-0.86), sensitivity=0.81 (95%CI=0.71-0.92), and specificity=0.67 (95%CI=0.55-0.78).

**Conclusions:** A panel of 55 blood leukocyte DNA methylation markers differentiates patients with metabolic HCC from control patients with benign metabolic liver disease, with a slightly higher sensitivity when combined with demographic and clinical information.

## Introduction

Metabolic-related liver disease is the fastest growing cause of liver cancer and its most common type, hepatocellular carcinoma (HCC)[1, 2]. Metabolic liver disease comprises metabolic dysfunction-associated steatotic liver disease (MASLD), and non-viral and non-alcoholic steatotic liver disease, and these are rapidly increasing worldwide [2–4]. Although chronic hepatitis B and C virus (HBV and HCV) infections were for several decades the major causes of HCC, improved treatment for HCV and increased vaccinations for HBV have shifted the burden of HCC to non-viral causes, with metabolic liver disease being the most rapidly rising cause [5]. Metabolic HCC exhibits unique molecular processes and immune characteristics and is considered a distinct HCC subtype, requiring characterization of its underlying molecular signatures, including epigenome-wide DNA methylation alterations [6].

Despite evidence of differential hepatotumorigenesis by cancer etiology [7, 8], most existing studies on DNA methylation profiles for HCC detection have been focused on all-cause HCC [9–13]. Few studies have assessed HCC detection in patients with viral hepatitis [14, 15] or all-cause liver cirrhosis [16, 17]. Genetically engineered mouse models with metabolic dysfunction-associated steatohepatitis (MASH)-related HCC suggest a distinct DNA methylation profile for the progression of MASH to HCC in murine models [18, 19]. Existing human studies on MASH-related HCC have focused on liver tissues, comparing DNA methylation status of tumor samples to paired adjacent noncancer tissues or to noncancer liver tissues from different individuals, but these have rarely been validated in circulating blood for noninvasive testing because of difficulty in obtaining appropriate patient samples [20, 21]. Identifying promising blood-based DNA methylation markers that could be combined with current clinical markers of HCC (e.g., alpha-feto protein [AFP], lectin-reactive AFP [AFP-L3], and des-γ-carboxy prothrombin [DCP]) would enhance clinical surveillance through noninvasive screening for metabolic HCC, a rapidly increasing global public health burden [1, 4].

The goal of this study was to perform an epigenome-wide DNA methylation profiling and validation in patients with metabolic HCC and in cancer-free control patients with metabolic liver disease in an international, multicenter study. Our primary aim was to identify differentially methylated 5’-C-phosphate-G-3’ (CpG) positions across the genome that discriminate metabolic HCC cases from metabolic controls. We also sought to develop and validate a multifactorial model combining CpGs with selected clinical and demographic variables. In secondary analysis, we assessed whether presence of the genetic risk variant *PNPLA3* (I148M) rs738409 could further improve metabolic HCC prediction by combining this variant with differentially methylated CpGs and clinical and demographic variables.

## Materials and Methods

### Study Population and Data Collection

Details of the design and methods used for participant recruitment and data collection have been published [22]. Briefly, data and biospecimen were obtained from the following six international sites: 1) the Barcelona Clinic Liver Cancer Group (BCLC), Hospital Clinic Barcelona and IDIBAPs, Barcelona, Spain; 2) Instituto de Investigación Sanitaria Biogipuzkoa (IISB), Donostia University Hospital, San Sebastian, Spain; 3) the Karolinska University Hospital, Sweden; 4) the Virgen del Rocio Hospital Institute of Biomedicine of Sevilla (IBIS), Seville, Spain; 5) the University of Texas Southwestern (UTSW), San Antonio, Texas; and 6) the Mayo Clinic sites in Rochester, Minnesota, and Jacksonville, Florida. All sites provided germline leukocyte DNA and epidemiological data on 673 metabolic HCC cases and 763 cancer-free controls with a history of MASLD (formerly known as nonalcoholic fatty liver disease [NAFLD]), metabolic syndrome, or other metabolic conditions (e.g., diabetes and obesity). Recruitment and data collection were completed at all sites before the recent change in the nomenclature from NAFLD to MASLD [3]. The participating sites were asked to exclude individuals with competing liver diseases before submitting their data and DNA samples to the Mayo Clinic. Potential participants excluded from the study included those with at least one of the following liver diseases: viral hepatitis (HBV, HCV), alcoholic liver disease, autoimmune hepatitis, alpha-1-antitrypsin deficiency, hemochromatosis, Wilson’s disease, biliary cirrhosis, primary sclerosing cholangitis, Budd-Chiari syndrome, and those who consumed ≥20 grams of alcohol per day. After these exclusions, metabolic HCC was defined as imaging or pathologically confirmed diagnosis of steatosis-related HCC, metabolic syndrome-related HCC, or cryptogenic HCC— most of which are associated with MASLD [23]. Controls were cancer-free individuals with imaging or pathological confirmation of hepatic steatosis. Data received from each site included information on case-control status, age at HCC diagnosis or recruitment for controls, sex, ethnicity, body mass index (BMI, kg/m^2^), smoking history, and type II diabetes mellitus status. For the present study, we frequency-matched 320 metabolic HCC cases with 320 metabolic controls based on age (± 5 years), sex, and study site for analyses. All participating sites obtained approval from their local institutional review boards (IRBs), and an additional IRB approval was obtained from the Mayo Clinic IRB for the present study (IRB#: 23-000005).

### DNA Methylation Assay and Quality Control Checks

Peripheral blood leukocyte DNA samples obtained from the participants were assayed on the Illumina Infinium Methylation EPIC BeadChip microarray (EPIC array; Illumina Inc., San Diego, CA, USA), which covers 850,000 CpG sites across the genome [24]. The assay was performed at the Mayo Clinic Genome Analysis Core laboratory. In brief, DNA quantification was performed using the Invitrogen Qubit dsDNA Quantification Assay kit (catalog# Q32853; ThermoFisher Scientific, Inc., Waltham, MA, USA). This was followed by a bisulfite modification process that utilized the column cleanup kit method under the alternative incubation conditions recommended by Illumina for the EPIC array. Measurements were done on a nanodrop instrument following the bisulfite modification. We ran the EPIC array using eight 96-well plates containing DNA from the 640 cases and controls. We included 16 laboratory control DNA samples (human methylated and unmethylated control DNA sets; catalogue #D5011 for methylated and #D5014 for unmethylated control DNA, Zymo Research Inc., Irvine, CA, USA). A pair of these methylated and unmethylated laboratory controls were included on each of the eight plates to determine if any of the probes should be excluded due to poor performance. Further, we included 64 participant duplicate samples that were distributed evenly across the plates. Determination of the methylation status of the target CpG sites involved comparing the ratio of a fluorescent signal from the methylated allele to the sum of the fluorescent signals from both methylated and unmethylated alleles (i.e., the β value). The β value per CpG range from 0 (unmethylated) to 1 (fully methylated). Both the laboratory controls and participant duplicates indicated excellent assay performance. The unmethylated laboratory controls showed an intraclass correlation of 0.95, while the methylated controls had a correlation of 0.83. For duplicates, we achieved correlations ≥0.98, and we retained the duplicated sample with the highest call rate in the final analysis. For further quality control (QC), CpGs were excluded if they were located at a single nucleotide polymorphism (SNP) location, failed in more than 10% of samples, were located on the X and Y chromosome, were determined to be cross-reactive, or overlapped with genetic variants [24].

This resulted in 691,187 CpGs passing QC. Data were normalized with *dasen* (*dasen* command in *watermelon* R package) that utilizes quantile normalization to normalize methylated and unmethylated intensities separately, and address types I and type II probes separately [25]. A small fraction of missing β values (<0.01%) were imputed using *champ.impute* function with k-nearest neighbor (KNN) and *k* parameter as five in the *ChAMP* R package. We used principal component analysis (PCA) to assess batch effect across the eight experimental plates. The PCA was performed on the top 2000 most variable autosome CpG probes, considering all samples (CpG probes with the largest standard deviations in M-values). We then used the Kruskal-Wallis rank-sum test to investigate the association between the top two principal components and the experimental plates, which did not show any association, ruling out batch effect as a concern. To account for differences in leukocyte cell types, we estimated cell type proportions for CD4 T cells (CD4T), CD8 T cells (CD8T), natural killer cells (NK-cells), B lymphocytes (B-cells), monocytes, and neutrophils using a customized set of probes obtained from IDOL optimization for blood as implemented in the *FlowSorted.Blood.EPIC* Bioconductor package [26]. For participant samples QC, we excluded samples with (1) poor assay performance based on the methylated and unmethylated intensity plot, (2) samples that failed biological sex check using the methods implemented in the *minfi* and *watermelon* R packages, and (3) samples determined to be outliers based on the *watermelon* method [25].

### Statistical Analysis

Differences in participant characteristics were compared using means and standard deviations (SDs) for continuous variables, and frequencies and percentages for categorical variables. The study sample was divided randomly into training (80%) and validation (20%) sets through a stratified approach based on frequencies that ensured approximately equal distributions by case-control status, age (5-year groups), sex, and study site in both the training and validation data. We assessed differences in the distribution of all study variables between the cases and controls in the training and validation data separately, but conclusions were based on results of the training data. The variables examined are age (continuous), sex, race (White, other), BMI (continuous), smoking history (never, former, current), diabetes mellitus (yes, no), study site (Mayo Clinic and UTSW combined, Karolinska hospital, BCLC-Barcelona and IISB-San Sebastian combined, and IBIS-Seville), and leukocyte cell type (CD4T, CD8T, NK-cells, B-cells, monocytes, and neutrophils). These comparisons were done using a Kruskal-Wallis rank-sum test for continuous variables and χ^2^ test for categorical variables. We combined data from UTSW with Mayo Clinic data because the UTSW data comprised only case subjects, and the IISB-San Sebastian data was combined with the BCLC-Barcelona data because the IISB data also comprised case subjects only. Variables found to be significantly different between cases and controls in the training data are race, diabetes mellitus, CD4T, monocytes, and neutrophils, and these were considered for further evaluation as covariates for (1) CpG selection (significant cell types), or (2) multifactorial prediction modeling (race and diabetes).

Candidate CpG selection and initial predictive modeling were done in the training data. Of the 691,187 CpGs that passed the QC checks, we used false-discovery rate (FDR)-corrected *p*-value (*q*-value), adjusting for the three significant cell types (CD4T, monocytes and neutrophils), to identify 164 differentially methylated CpGs that met the significance threshold (*q*<0.05) (**Suppl. Table 1**). These CpGs were identified by comparing the metabolic HCC cases with the metabolic controls in the training data and using the moderated paired *t*-test from the R Bioconductor package, linear models for microarray data (*limma*) [27]. To address high-dimensionality and multicollinearity among the selected CpGs, LASSO regression with 10-fold cross validation was employed using a generalized linear model via penalized maximum likelihood (*glmnet*). The grid search in *glmnet* involved keeping the alpha value fixed at one and varying lambda (regularization parameter) values. The LASSO regression process generated shrunken estimates for each CpG, and we retained only 55 CpGs with nonzero coefficients for prediction modeling, as these are the most informative markers. We used a Manhattan plot to visualize the CpGs across chromosomes, and a volcano plot to visualize the hypomethylated and hypermethylated CpGs. Methylation values of the CpGs were also visualized using heatmaps. These data visualizations were done using the R packages *ggplot2* and *ComplexHeatmap*.

In our primary analysis, we first constructed a predictive model that included key biological variables (age and sex), and the significant demographic and clinical variables described above (race and diabetes) using area under the receiver operating characteristic curve (AUC-ROC) analysis with the R package *pROC.* This initial model was constructed to provide a baseline context for evaluating the predictive value of the identified CpGs. We followed this with a predictive model that included only the parsimonious list of 55 differentially methylated CpGs using AUC-ROC analysis. We then constructed a multifactorial model that combined the key biological-demographic and clinical variables (age, sex, race, diabetes) with the 55 differentially methylated CpGs in the same model to evaluate the performance of an elaborate model and compare with the performance of the CpGs only model. We performed two secondary analyses. In the first secondary analysis, we evaluated the additional predictive impact of *PNPLA3*-rs738409 in a subgroup of participants with genetic data available from our previous study [22]. Here too, we constructed a base model that included only the demographic and clinical variables (age, sex, race, diabetes mellitus) and rs738409 using AUC-ROC analysis. This was also followed by a separate model for only the 55 CpGs in this subgroup of participants. We then constructed an elaborate model that included age, sex, race, diabetes mellitus, rs738409, and the 55 CpGs. All training data predictive models underwent validation in an independent 20% of the sample, evaluating AUC, sensitivity, and specificity. In the second secondary analysis, we built similar predictive models using only the hypermethylated CpGs. The underlying methylation data and limited covariates have been made available in NCBI/GEO: ID# GSE281691.

## Results

Of the 640 participant samples included in the study, one sample showed poor assay performance, 46 samples had discordance between self-reported sex and biological sex inferred from the X:Y chromosome, and five samples were identified as outliers. After excluding these samples, 588 samples remained for analyses (272 metabolic HCC cases and 316 metabolic controls) (**Table 1**). Briefly, in the overall sample, there was a greater representation of men (65%), non-Hispanic Whites (87%), and individuals with type II diabetes mellitus (65%). Data on the *PNPLA3*-rs738409 genetic risk variant was available on 75% (n=439) of participants. We split the overall sample in 80:20 ratio into training (n=469) and validation (n=119) sets. The cases and controls did not differ significantly by age, sex, or study site in either the training or validation sample (**Table 1**). In the training data, cases had greater proportions of non-Whites and individuals with a history of type II diabetes mellitus than controls. The case participants had also higher leukocyte proportions of CD4T, monocytes, and neutrophils than did the controls. There were no other significant differences observed in the training data. In the validation data, only non-Whites, individuals with diabetes mellitus, and those with a higher monocyte cell type proportions were higher in cases than controls.

**Table 1.**
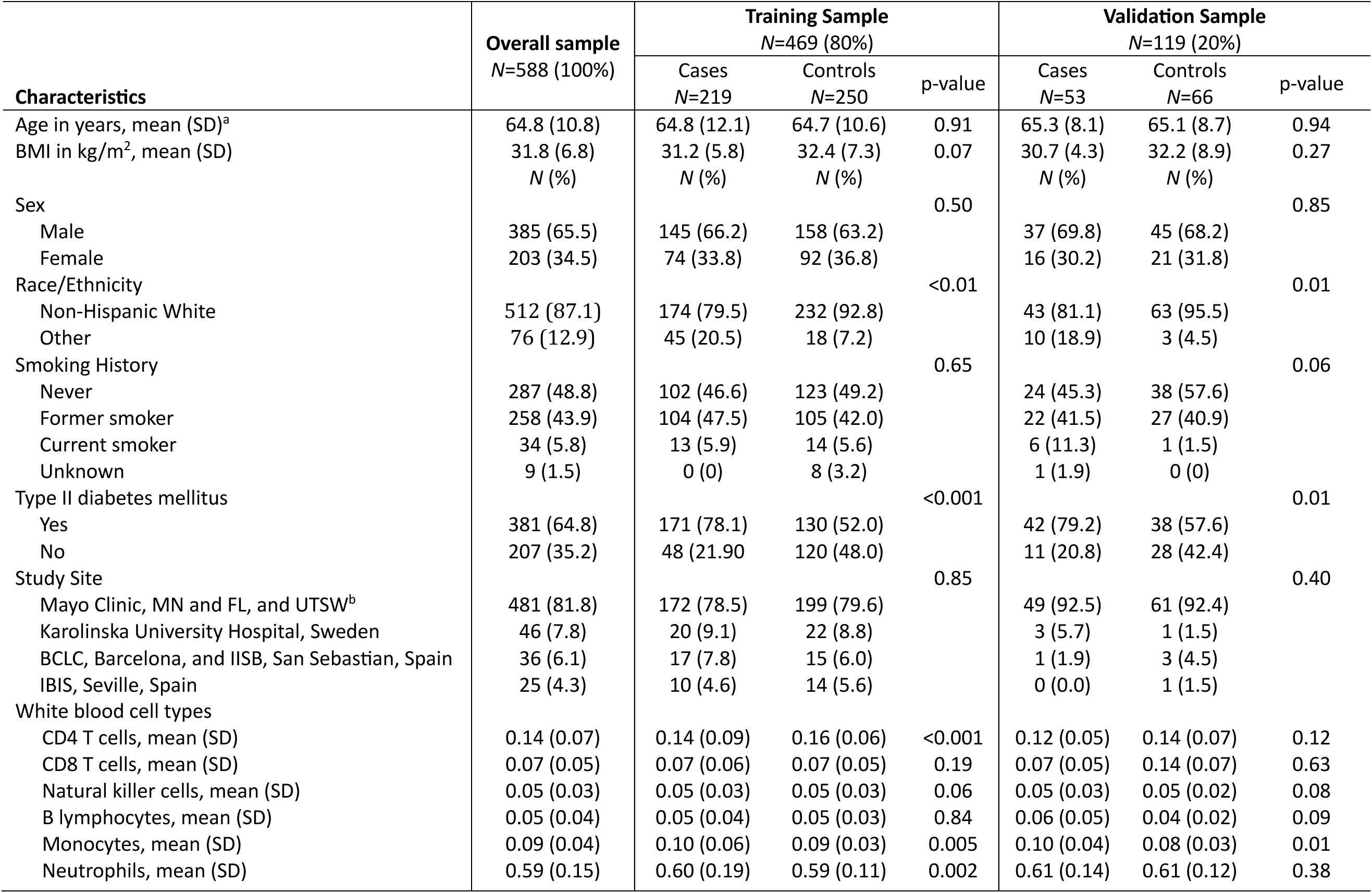
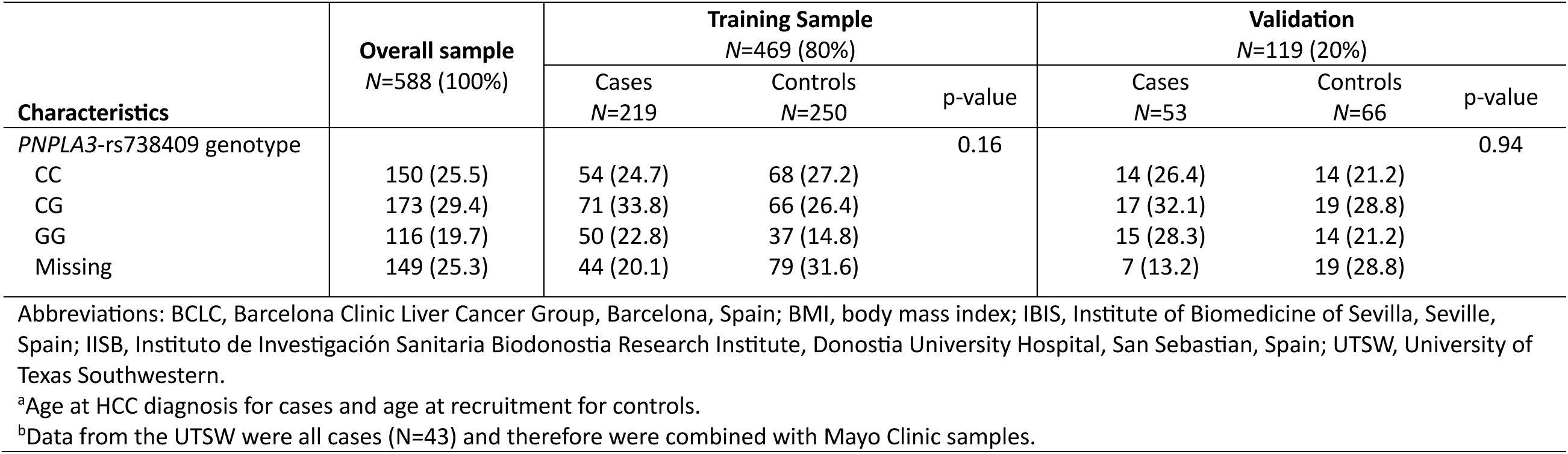
Descriptive Statistics of the Training and Validation Samples.

We performed an epigenome-wide association study (EWAS) in the training data based on 691,187 CpGs that passed QC (**Figures 1A-1D**). The EWAS did not show overfitting as the genomic inflation lambda value is closer to one, which is within the expected range (λ=1.31, **Figure 1B**). Of the 691,187 CpG sites, 164 were differentially methylated (110 hypermethylated and 54 hypomethylated) in the metabolic HCC cases compared to metabolic controls (**Figures 1A** and **1C**, and **Suppl. Table 1**). We used LASSO regression with 10-fold cross validation to assess multicollinearity and reduced the EWAS significant CpGs to a parsimonious list of 55 informative markers with non-zero coefficients, of which 33 were hypermethylated and 22 were hypomethylated (**Figure 1D** and **Table 2**). Interestingly, many of the genes linked to the differentially methylated CpGs have been associated with liver disease progression (e.g., *DCP2, TRPV3, ARRB1, KCNIP4, MIR10A*), and cancer formation or progression (e.g., *MTHFR, GRIK2, GSN, HOX3, KCNMA1*) (**Table 2**).

**Figure 1.**
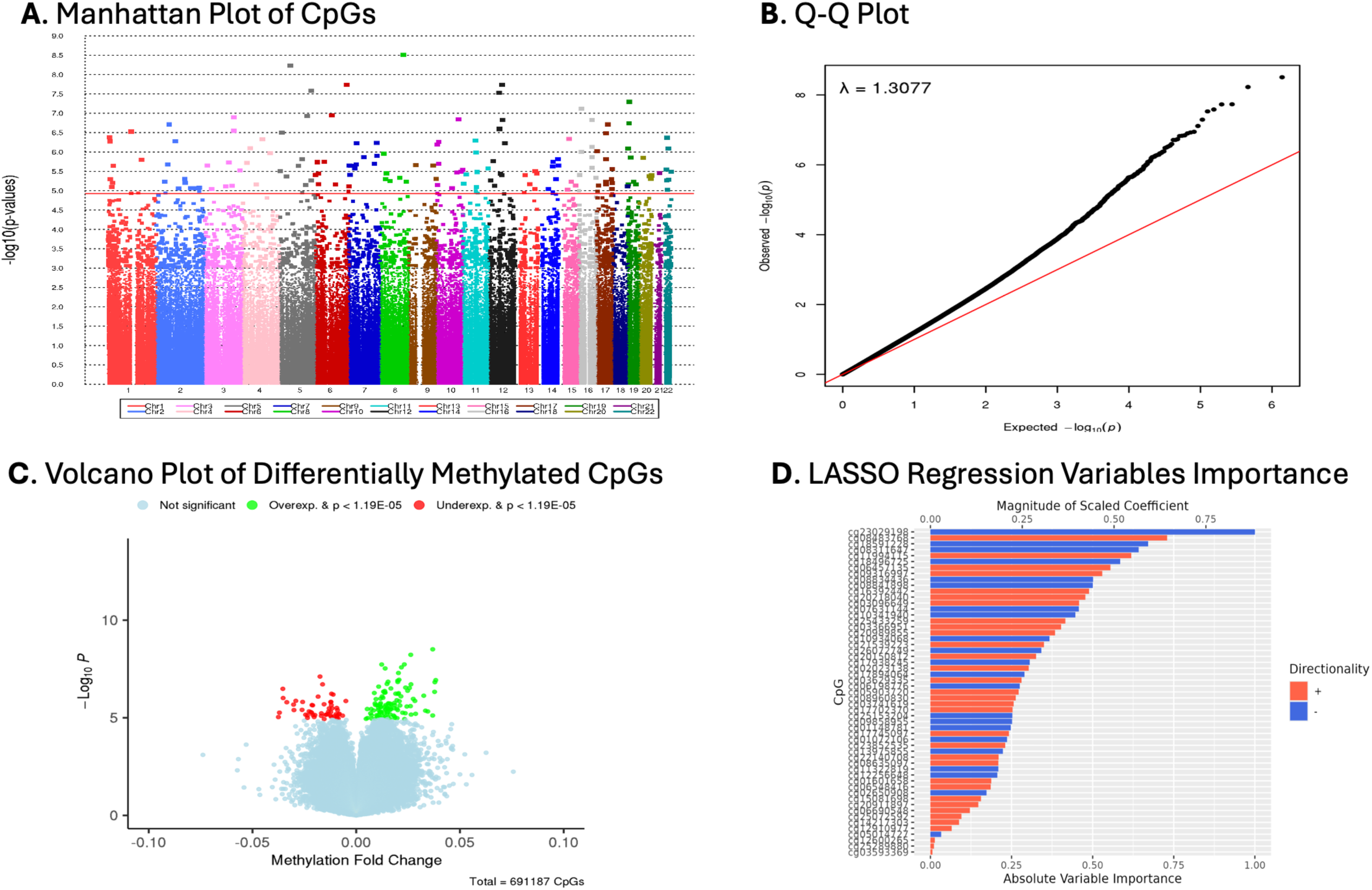
Epigenome-wide analysis for selection of differentially methylated CpGs associated with metabolic HCC. The analysis was performed among 272 Metabolic HCC cases and 316 metabolic controls. (**A**) Manhattan plot with false discovery rate (FDR)-adjusted *p*-value threshold (red horizontal line) for selection of significant CpGs (*q*-value<0.05; n=164 CpGs) in the training data for further screening. (**B**) Q-Q plot of CpGs showing a lambda (λ) value that is closer to 1. (**C**) Volcano plot of the 164 FDR-significant CpGs, showing hypomethylated CpGs in red color and hypermethylated CpGs in green color among cases versus controls in the training data. (**D**) Results of a LASSO regression model with 10-fold cross validation, reducing the 164 FDR-significant CpGs to a parsimonious list of 55 CpGs with non-zero coefficients (33 hypermethylated and 22 hypomethylated) and scaling of absolute importance of each CpG in the presence of the other CpGs. This is the final set of CpGs used for the primary analysis.

**Table 2.** Differentially methylated CpGs used for the primary analysis; *N*=55 CpGs (33 hypermethylated and 22 hypomethylated).

| CpG Probe | Chromosomal position<sup>a</sup> | Genes | Relation to Island | Coefficient | Cases: Mean beta value | Controls: Mean beta value | Log-fold change<sup>b</sup> | Raw P-value |
| --- | --- | --- | --- | --- | --- | --- | --- | --- |
| cg25433259 | Chr6:159271784 | *OSTCP1* | OpenSea | 5.1082312 | 0.66541858 | 0.63835286 | 0.01226212 | 1.86E-08 |
| cg16392442 | Chr12:49595954 | - | OpenSea | 15.2001542 | 0.17353326 | 0.15696093 | 0.01394544 | 2.95E-08 |
| cg06457135 | chr19:5953406 | *RANBP3* | OpenSea | 9.3450824 | 0.40866314 | 0.38436786 | 0.02035666 | 5.09E-08 |
| cg12910977 | chr5:138207492 | *LRRTM2;CTNNA1* | OpenSea | 0.7322945 | 0.56139306 | 0.5179095 | 0.03832187 | 1.18E-07 |
| cg02023138 | chr12:51701979 | *BIN2* | OpenSea | 11.3546081 | 0.84181837 | 0.83196508 | 0.01081285 | 2.53E-07 |
| cg07631144 | chr17:46657393 | *MIR10A* | N_Shore | -4.1056162 | 0.29359393 | 0.33761632 | -0.0353619 | 3.28E-07 |
| cg03629335 | chr4:100126967 | *ADH6* | OpenSea | 2.8296323 | 0.69035821 | 0.65791421 | 0.03776824 | 4.69E-07 |
| cg17745097 | chr1:11863365 | *MTHFR* | N_Shelf | 10.4633105 | 0.08307216 | 0.07659769 | 0.00870919 | 5.36E-07 |
| cg17938245 | chr12:80067352 | *PAWR* | OpenSea | -5.5211165 | 0.68665745 | 0.693618 | -0.0119055 | 5.92E-07 |
| cg20911897 | chr4:37953726 | *TBC1D1* | OpenSea | 1.9160149 | 0.46867666 | 0.45262885 | 0.02157469 | 7.96E-07 |
| cg06690548 | chr4:139162808 | *SLC7A11* | OpenSea | 1.8024676 | 0.78789942 | 0.76138798 | 0.0274857 | 1.07E-06 |
| cg11322819 | chr19:13694039 | - | OpenSea | -14.1189425 | 0.07830778 | 0.08513437 | -0.0049903 | 1.38E-06 |
| cg17894064 | chr5:112657195 | *MCC* | OpenSea | -4.1526787 | 0.73211569 | 0.7569463 | -0.0258219 | 1.54E-06 |
| cg22140708 | chr6:10081664 | - | OpenSea | 5.1449817 | 0.78206787 | 0.77170398 | 0.0154948 | 1.83E-06 |
| cg20989855 | chr4:20985927 | *KCNIP4* | OpenSea | 9.0390939 | 0.22096169 | 0.20473515 | 0.0164293 | 1.90E-06 |
| cg08311647 | chr10:11596182 | *USP6NL* | OpenSea | -8.2102209 | 0.32034876 | 0.33414088 | -0.0155017 | 2.03E-06 |
| cg03096649 | chr7:130056977 | *CEP41* | OpenSea | 13.3323684 | 0.80181661 | 0.78461484 | 0.01153379 | 2.03E-06 |
| cg09858955 | chr2:58135951 | *VRK2* | OpenSea | -2.8610325 | 0.35796606 | 0.3978885 | -0.0302066 | 2.11E-06 |
| cg05903720 | chr14:104663241 | - | OpenSea | 5.7993613 | 0.67104233 | 0.65604405 | 0.01851333 | 2.23E-06 |
| cg20150812 | chr9:124029753 | *GSN* | OpenSea | 5.1334782 | 0.63156105 | 0.6199896 | 0.01418724 | 2.25E-06 |
| cg25072592 | chr14:75355586 | *DLST* | OpenSea | 1.5676053 | 0.53199877 | 0.51258117 | 0.0210767 | 2.34E-06 |
| cg17702370 | chr17:79283128 | *C17orf55* | N_Shore | 8.2558677 | 0.15774941 | 0.14859067 | 0.0119348 | 2.79E-06 |
| cg20218040 | chr6:14369697 | - | OpenSea | 8.2222083 | 0.78452904 | 0.76199551 | 0.01677532 | 3.60E-06 |
| cg12600265 | chr7:5422883 | *TNRC18* | OpenSea | 0.2650506 | 0.57937612 | 0.56715465 | 0.01639226 | 3.88E-06 |
| cg10934068 | chr6:509762 | *EXOC2* | OpenSea | -3.3359868 | 0.58562148 | 0.59379713 | -0.0261018 | 3.92E-06 |
| cg01148781 | chr8:99963021 | *OSR2* | S_Shore | -4.0754728 | 0.127759 | 0.14846004 | -0.0227403 | 4.70E-06 |
| cg08483768 | chr16:86304619 | - | OpenSea | 8.0038342 | 0.50945873 | 0.48237272 | 0.0344807 | 4.72E-06 |
| cg01072106 | chr9:138952311 | *NACC2* | OpenSea | -5.4605017 | 0.66997388 | 0.69090851 | -0.0133822 | 4.89E-06 |
| cg09316997 | chr14:91874913 | *CCDC88C* | OpenSea | 6.4844744 | 0.58855649 | 0.56312738 | 0.01150639 | 5.05E-06 |
| cg02650908 | chr17:74889830 | *MGAT5B* | OpenSea | -3.0128946 | 0.7169462 | 0.73871776 | -0.0218802 | 5.07E-06 |

**Table 2** (continued).
| CpG Probe | Chromosomal position<sup>a</sup> | Gene | Relation to Island | Coefficient | Cases: Mean beta value | Controls: Mean beta value | Log-fold change<sup>b</sup> | Raw *P*-value |
| --- | --- | --- | --- | --- | --- | --- | --- | --- |
| cg25289880 | chr1:13856326 | - | OpenSea | 0.3631003 | 0.11750793 | 0.10623291 | 0.01184671 | 5.07E-06 |
| cg08834436 | chr22:27831818 | - | N_Shelf | -5.1226004 | 0.33084528 | 0.36666469 | -0.0370071 | 5.43E-06 |
| cg03593369 | chr2:43456557 | - | S_Shore | 0.1501873 | 0.29323094 | 0.27353325 | 0.01358761 | 5.89E-06 |
| cg13975855 | chr17:46652550 | *HOXB3* | N_Shore | -4.3014449 | 0.7616477 | 0.78337037 | -0.0167974 | 5.93E-06 |
| cg06548416 | chr1:24438703 | *MYOM3* | OpenSea | 2.4685847 | 0.23242444 | 0.20555792 | 0.02863055 | 6.24E-06 |
| cg23029198 | chr2:146510791 | - | OpenSea | -39.0402834 | 0.79023973 | 0.79559585 | -0.0081623 | 6.25E-06 |
| cg18591228 | chr11:3175552 | *OSBPL5* | OpenSea | -8.1292725 | 0.35349018 | 0.37448402 | -0.0097212 | 6.57E-06 |
| cg06198776 | chr13:73557424 | *PIBF1* | OpenSea | -11.514518 | 0.87081259 | 0.88156024 | -0.008382 | 6.73E-06 |
| cg26072749 | chr17:46657274 | *MIR10A* | N_Shore | -6.10247 | 0.15782119 | 0.17616999 | -0.0143176 | 6.87E-06 |
| cg11994115 | chr19:40360856 | *FCGBP* | N_Shore | 8.7958448 | 0.64306667 | 0.61853271 | 0.02644958 | 6.87E-06 |
| cg15081698 | chr6:101847050 | *GRIK2* | Island | 5.0372892 | 0.12331257 | 0.11170805 | 0.01080277 | 6.97E-06 |
| cg18496725 | chr4:68788615 | *TMPRSS11A* | OpenSea | -9.0086014 | 0.75106914 | 0.77602459 | -0.0244003 | 6.97E-06 |
| cg05014727 | chr10:6214016 | *PFKFB3* | OpenSea | -0.373298 | 0.28626944 | 0.31704808 | -0.0198905 | 7.25E-06 |
| cg23852535 | chr17:8857258 | *PIK3R5* | OpenSea | 10.6771863 | 0.83787502 | 0.83078653 | 0.00737971 | 7.32E-06 |
| cg14217303 | chr15:85177537 | *SCAND2P* | S_Shore | 3.2816993 | 0.25645962 | 0.24704736 | 0.00983248 | 7.36E-06 |
| cg12256648 | chr3:143752097 | - | OpenSea | -3.2169829 | 0.42099641 | 0.44279351 | -0.0234836 | 7.53E-06 |
| cg08841898 | chr12:27717865 | *PPFIBP1* | OpenSea | -18.5292437 | 0.68745191 | 0.70022304 | -0.0092687 | 7.55E-06 |
| cg10341940 | chr18:76822780 | - | OpenSea | -24.5248737 | 0.82390723 | 0.82977233 | -0.006565 | 7.77E-06 |
| cg25153204 | chr10:79291246 | *KCNMA1* | OpenSea | -7.5939005 | 0.84999289 | 0.85835243 | -0.0110492 | 8.58E-06 |
| cg03366951 | chr3:39302545 | - | OpenSea | 6.709593 | 0.16306437 | 0.15399861 | 0.01408014 | 8.95E-06 |
| cg03741619 | chr17:3438918 | *TRPV3* | Island | 16.8968122 | 0.07389002 | 0.06818281 | 0.00544581 | 1.03E-05 |
| cg01601658 | chr6:168785524 | - | OpenSea | 3.3166661 | 0.30778145 | 0.288747 | 0.01968328 | 1.04E-05 |
| cg08635097 | chr13:44833857 | - | OpenSea | 9.6920045 | 0.12942612 | 0.12314091 | 0.00755129 | 1.11E-05 |
| cg21539223 | chr5:112312093 | *DCP2* | N_Shore | 27.1214227 | 0.04927896 | 0.04464676 | 0.00476079 | 1.13E-05 |
| cg08960830 | chr11:75047180 | *ARRB1* | OpenSea | 8.8896234 | 0.69990481 | 0.69248749 | 0.00905003 | 1.16E-05 |

<sup>a</sup>Positions are based on the human reference genome assembly GRCh38.<sup>b</sup>Fold change comparing beta estimates between cases and controls.

To provide a context for assessing the discriminatory accuracy of the 55 informative CpGs, we first created a base model comprising demographic and clinical variables only: age, sex, race, and diabetes mellitus. This base model yielded a training sample AUC=0.66 (95% CI: 0.61-0.71), sensitivity=0.81 (95% CI: 0.76-0.86), and specificity=0.47 (95% CI: 0.41-0.53), and validation sample AUC=0.65 (95% CI: 0.55-0.75), sensitivity=0.62 (95% CI: 0.49-0.75), and specificity=0.64 (95% CI: 0.52-0.75) (**Figure 2A)**. Next, we assessed the predictive accuracy of only the parsimonious panel of 55 informative CpGs in the training data, yielding AUC=0.97 (95% CI: 0.96-0.99), sensitivity=0.93 (95% CI: 0.89-0.96), and specificity=0.93 (95% CI: 0.90-0.96) (**Figure 2B**). The validation results for the CpGs only model was AUC=0.79 (95% CI: 0.71-0.87), sensitivity=0.77 (95% CI: 0.66-0.89), and specificity=0.74 (95% CI: 0.64-0.85) (**Figure 2B**). We then assessed the combined predictive ability of an elaborate model that included age, sex, race, diabetes mellitus, and the 55 CpGs, yielding training sample AUC=0.98 (95% CI: 0.97-0.99), sensitivity=0.92 (95% CI: 0.89-0.96), and specificity=0.96 (95% CI: 0.94-0.96). Results from the validation sample for the joint elaborate model were AUC=0.78 (95% CI: 0.70-0.86), sensitivity=0.81 (95% CI: 0.71-0.92), and specificity=0.67 (95% CI: 0.55-0.78) (**Figure 2C**). These results constitute our primary findings.

**Figure 2.**
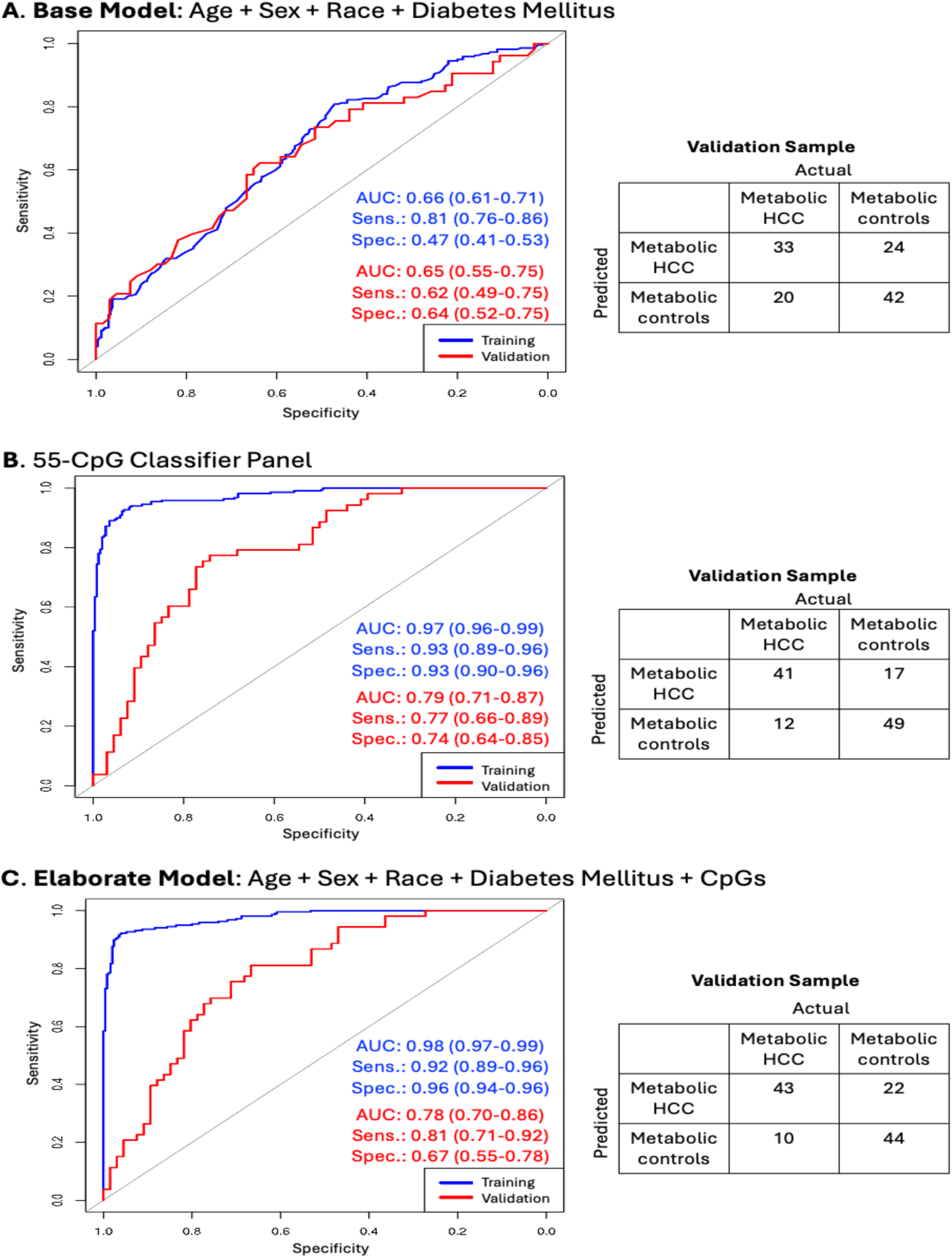
Distinguishing metabolic HCC from benign metabolic liver disease using demographic and clinical variables and differentially methylated CpGs. The study sample comprised 272 Metabolic HCC cases and 316 metabolic controls. (**A**) Training and validation results from area under the receiver operating characteristic curve (AUC-ROC) analysis for a model that included age (continuous), sex, race (White, other), and type II diabetes mellitus (yes, no). (**B**) AUC-ROC analysis for a model that included only the 55 differentially methylated CpGs as shown in Table 2. (**C**) An elaborate multifactorial AUC-ROC analysis for a model that included age, sex, race, diabetes mellitus. and the 55 CpGs. Abbreviations: AUC, area under the receiver operating curve; HCC, hepatocellular carcinoma; sens., sensitivity; spec.: specificity.

In secondary analysis among a subgroup of participants with genetic data, we assessed the additional predictive impact of the HCC susceptibility variant, *PNPLA3*-rs738409 (**Figure 3**). Here too, we created a base model that comprised only age, sex, race, diabetes mellitus and rs738409, yielding validation sample AUC=0.66 (95% CI: 0.54-0.77), sensitivity=0.80 (95% CI: 0.69-0.92), specificity=0.45 (95% CI: 0.30-0.59) (**Figure 3A**). Validation results for a model with only the 55 CpGs in this subgroup were AUC=0.76 (95% CI: 0.66-0.86), sensitivity=0.76 (95% CI: 0.64-0.88), and specificity=0.70 (95% CI: 0.57-0.83) (**Figure 3B**). Further, we built an elaborate model that assessed the combined predictive ability of the clinical, demographic, and genetic data together with the 55 CpGs in the subgroup of participants with available genetic data. After running a penalized LASSO regression analysis for the elaborate model in the subgroup analysis, only 44 of the CpGs had non-zero coefficients (**Suppl. Table 2**), together with age, sex, race, diabetes mellitus and rs738409 were used for prediction modeling. This elaborate model yielded a validation AUC=0.75 (95% CI: 0.65-0.85), sensitivity=0.74 (95% CI: 0.61-0.87), and specificity=0.70 (95% CI: 0.57-0.83) (**Figure 3C**).

**Figure 3.**
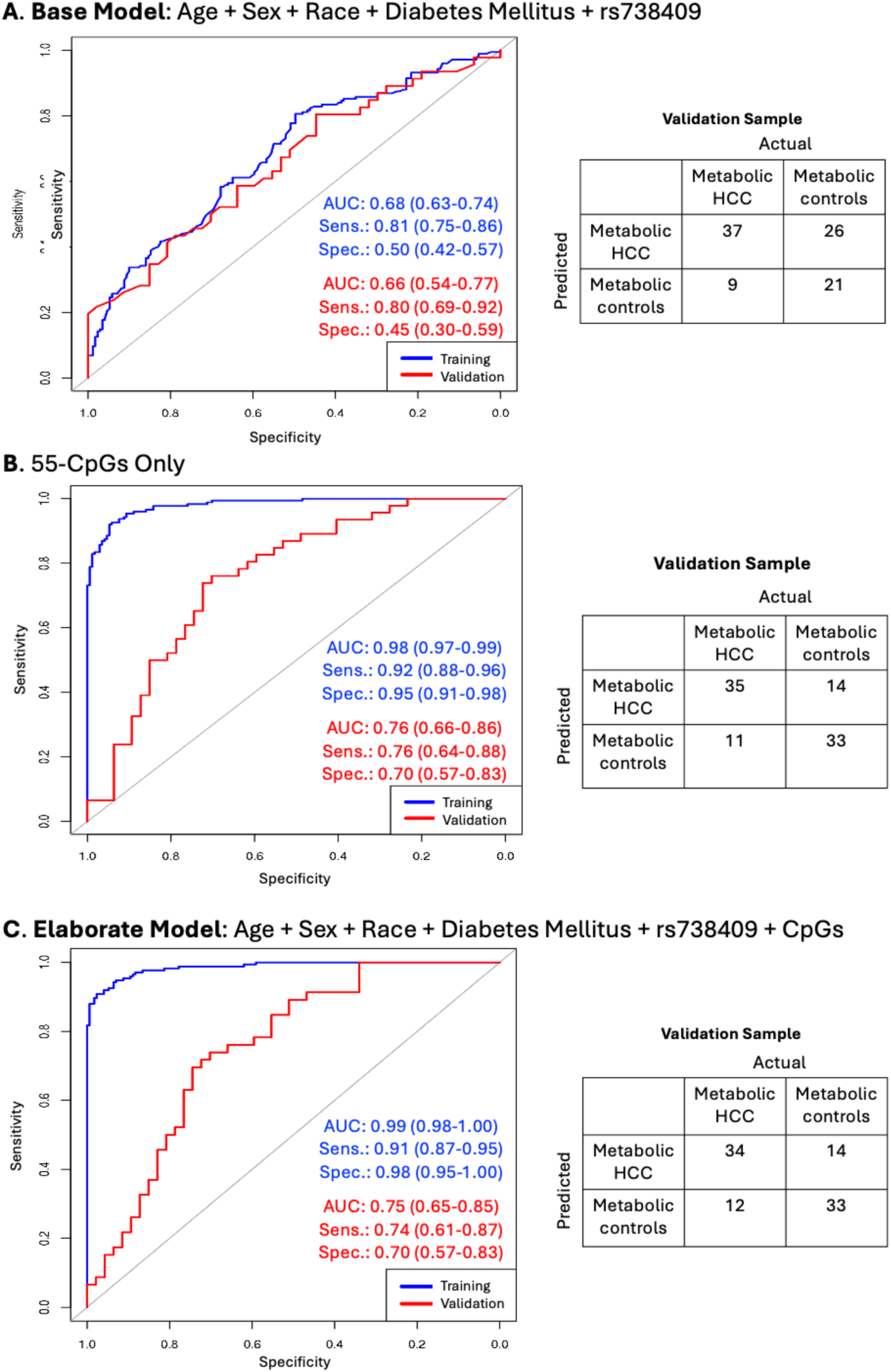
Discriminating between metabolic HCC and metabolic liver disease in a subgroup of participants with genetic data. These analyses were performed among 75% of the study sample (n=439). (**A**) Training and validation results from area under the receiver operating characteristic curve (AUC-ROC) analysis for a model that included age (continuous), sex, race (White, other), diabetes mellitus (yes, no), and *PNPLA3*-rs738409 genotype. (**B**) Training and validation results for a model that included only the 55 differentially methylated CpGs as shown in Table 2. (**C**) Multifactorial AUC-ROC analysis for metabolic HCC combining the clinical and demographic variables with CpGs. This multifactorial model was built using LASSO regression with 10-fold cross validation and examining the clinical and demographic variables and the 55 CpGs. However, only 44 CpGs with non-zero coefficients were retained in addition to age, sex, race, diabetes mellitus, and rs738409 for prediction modeling. Abbreviations: AUC, area under the receiver operating curve; HCC, hepatocellular carcinoma; sens., sensitivity; spec.: specificity.

We repeated all analyses using only the hypermethylated CpGs from the EWAS significant CpGs (n=110, *q*<0.05). Based on a penalized LASSO regression analysis with 10-fold cross validation, we identified a 42-CpG classifier panel with non-zero coefficients that showed differential methylation values between cases and controls (**Figure 4A-4C** and **Suppl. Table 3**). Upon fitting the 42 hypermethylated CpGs, we observed validation AUC=0.75 (95% CI: 0.66-0.84), sensitivity=0.81 (95% CI: 0.71-0.92), and specificity=0.62 (95% CI: 0.50-0.74) (**Figure 4D**). We performed a separate multifactorial penalized LASSO regression analysis that included age, sex, race, diabetes mellitus, and the 42 CpGs, retaining 40 CpGs with non-zero coefficients (**Suppl. Table 4**) together with age, sex, race, and diabetes mellitus. This yielded a validation AUC=0.75 (95% CI: 0.66-0.84), sensitivity=0.72 (95% CI: 0.60-0.84), and specificity=0.73 (95% CI: 0.62-0.83) (**Figure 4E**). We further constructed an independent model in the subgroup of participants with genetic data, fitting a penalized LASSO regression analysis with the 42 hypermethylated CpGs, retaining 38 CpGs (**Suppl. Table 5**) together with age, sex, race, diabetes mellitus and rs738409 for prediction modeling. This resulted in a validation AUC=0.75 (95% CI: 0.65-0.85), sensitivity=0.83 (95% CI: 0.72-0.94), and specificity=0.62 (95% CI: 0.48-0.76) (**Figure 4F**).

**Figure 4.**
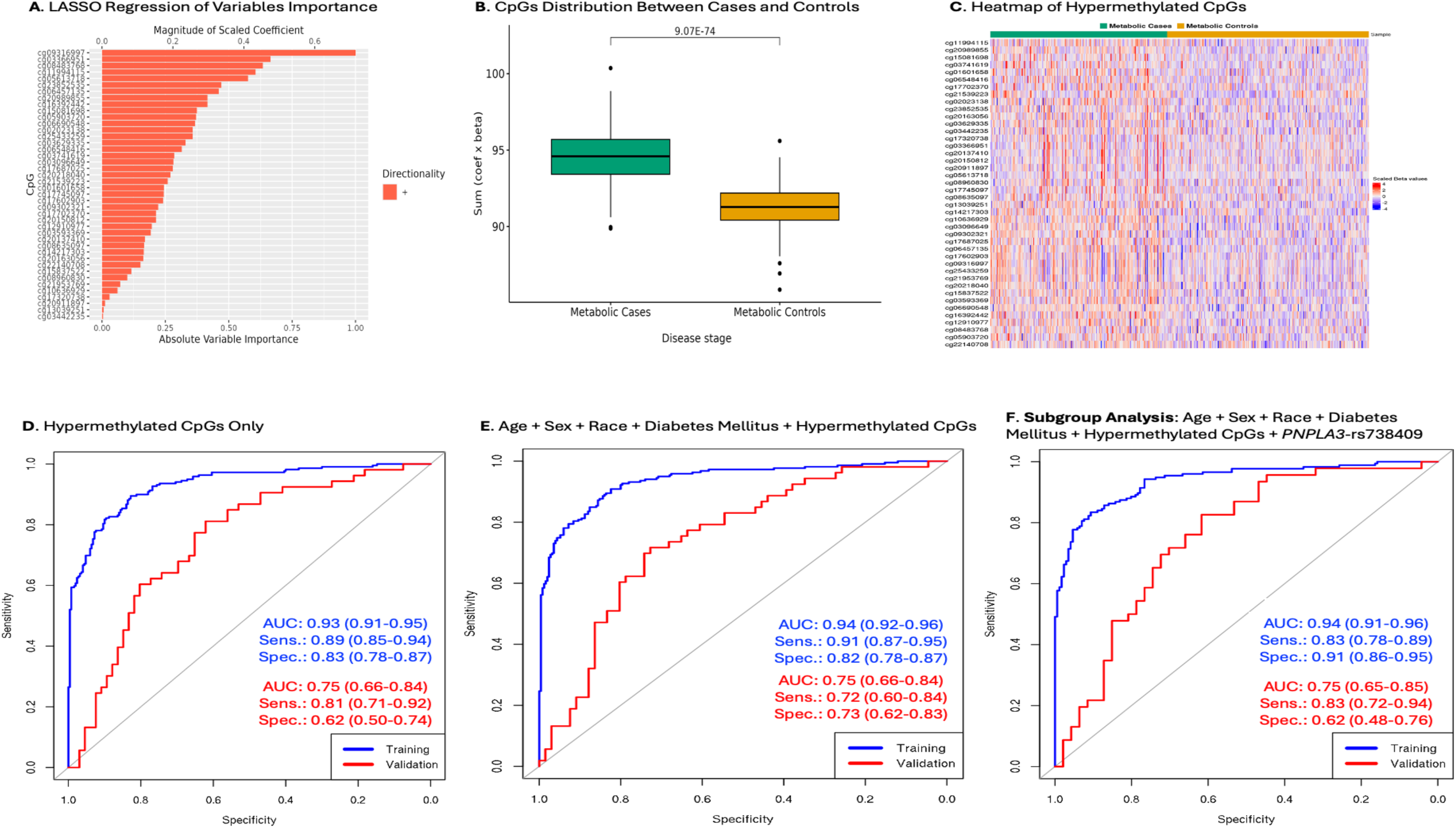
Characterizing metabolic HCC using hypermethylated CpGs only, and in combination with clinical, demographic, and *PNPLA3*-rs738409. The analysis was performed among 272 Metabolic HCC cases and 316 metabolic controls. (A) LASSO regression with scaled absolute importance of 42 hypermethylated CpGs used for the CpGs only model. (B) Differential distribution of the combined product of the 42 hypermethylated CpGs (estimated coefficients x beta values) between cases and controls. (C) Heatmap of 42 selected CpGs in the training data. (D) Modeling of area under the receiver operating characteristic curves (AUC-ROCs) for the hypermethylated CpGs only (n=42) in the training and validation samples. (E) A separate model that evaluated the combination of age (continuous), sex, race (White, other), type II diabetes mellitus (yes, no), and the hypermethylated CpGs in a distinct LASSO regression model with 10-fold cross validation, retaining 40 hypermethylated CpGs plus age, sex, race, and diabetes for prediction modeling. (F) A subgroup analysis modeling AUCs for the hypermethylated CpGs plus age, sex, race, diabetes, and *PNPLA3*-rs738409 among participants with genetic data (n=439) using a separate LASSO regression with 10-fold cross validation. This analysis retained 38 CpGs, age, sex, race, diabetes, and rs738409 for prediction modeling in the training (n=346) and validation (n=93) samples. Abbreviations: AUC, area under the receiver operating curve; HCC, hepatocellular carcinoma; sens., sensitivity; spec.: specificity.

## Discussion

In this large multicenter study, we performed an EWAS in patients with metabolic liver disease from which 55 differentially methylated CpGs were identified and independently validated for association with metabolic HCC. To provide a context for evaluating the predictive accuracy of the identified CpGs, we first constructed a base model that comprised age, sex, race and diabetes mellitus, yielding validation AUC of 0.65, sensitivity of 0.62 and specificity of 0.64, and this base model did not perform as well as our 55-CpG classifier model with validation AUC of 0.79, sensitivity of 0.77, and specificity of 0.74. We also developed a multifactorial model that combined age, sex, race, and diabetes mellitus with the 55-CpG panel, and this elaborate model had slightly higher sensitivity but lower specificity in the validation sample (AUC=0.78, sensitivity=0.81, specificity=0.67) compared to the 55-CpGs only model. Further, we explored a multifactorial model in a subgroup of participants with genetic data, jointly assessing the predictive accuracy of age, sex, race, diabetes mellitus, *PNPLA3*-rs738409, and the 55 CpGs. Validation results of this model (AUC=0.75, sensitivity=0.74, specificity=0.70) did not differ substantially from a model built with only the 55 CpGs in the same subgroup of participants (AUC=0.76, sensitivity=0.76, specificity=0.70). Together, the sensitivity values of these models are higher or nearly at par with reported sensitivity of AFP, the most widely used HCC diagnostic marker, with published sensitivity values of AFP ranging between 0.48 and 0.84 for the detection of all-cause HCC [14, 15]. However, because 20-30% of HCC tumors do not secrete AFP [28], future studies that combine relevant CpGs with AFP or other clinical diagnostic markers (e.g., DCP) and genetic risk variants for multifactorial modeling could enhance prediction of metabolic HCC in patients with metabolic liver disease.

DNA methylation plays an important role in transcriptome regulation and gene expression [29]. Aberrant DNA methylation has been found to be stably maintained by the DNA methyltransferase genes, *DNMT1, DNMT3A* and *DNMT3B,* during multistage tumorigenesis of various malignancies [21, 29]. Tumor suppressor gene silencing through DNA hypermethylation and oncogene activation through DNA hypomethylation can both contribute to cancer development, and these methylation markers could become potential targets of therapy [30, 31]. In hepatic tumorigenesis, aberrant DNA methylation has been observed in the development of HCC, but most methylation studies have focused on single gene loci [32] or a target candidate gene panel [9, 33, 34] and even all-cause HCC [9–13], but these have not proven to be sufficiently robust when compared to AFP and other clinical diagnostic biomarkers. A meta-analysis of 20 studies on all-cause HCC found that target candidate gene-based CpG panels do not perform adequately well to inform clinical test development [35]. Our use of an unbiased EWAS approach for screening of informative markers has the advantage of identifying potentially novel methylation markers for etiology-specific HCC detection, which is important for metabolic HCC given evidence of its distinct molecular signatures [6], and its fast-rising incidence worldwide [1, 2].

Because aberrant methylation can repress tumor suppressor genes or enhance oncogene activity [30, 31], it is important to assess the effects of both hypermethylated and hypomethylated CpGs jointly regarding tumor development. In our primary analysis, we identified 33 hypermethylated and 22 hypomethylated CpGs that play potential roles in metabolic HCC development (**Table 2**). Among the genes linked to hypermethylation in the cases, *TRPV3* [36], *DCP2* [37], *KCNIP4* [38], and *ARRB1* [39] have been associated with progression of liver disease to fibrosis and cirrhosis. Upregulation of *ARRB1* has been further found to induce inflammation-associated HCC development, while inhibition of this gene reduces hepatic inflammation and hepatotumorigenesis [39]. Other studies have found higher expression of *ARRB1* during HCC metastasis [40], and its upregulation correlates with tumor progression [41]. *MTHFR* is also one of the hypermethylated CpG-linked genes found in this study, and polymorphisms in this gene, which is involved in one-carbon metabolism of folate, have been associated with higher HCC risk and poor prognosis of HCC patients [42, 43]. Further, *GRIK2* has been associated with liver cancer development and metastasis [44]. *In vivo* experiments have also shown that overexpression of *GSN,* another hypermethylated CpG-linked gene, promotes HCC development through inhibition of the *TP53* tumor suppressor gene [45]. Moreover, *GSN* has been found to promote HCC invasion and metastasis through its regulation of epithelial-mesenchymal transition [46, 47].

Among the hypomethylated genes, a study by Wang *et al.* suggests that *HOXB3* is downregulated in cryptogenic HCC development [48]. *HOXB3*, which is involved in several cellular processes, including cell growth and differentiation, has also been found to be downregulated in breast and pancreatic cancers [49]. In another study, *HOXB3* was found to interact with *DNMT3B* to promote leukemia development [50]. *MIR10A* has been proposed as a marker for liver fibrosis development in chronic liver disease [51] and has also been found to promote HCC cell proliferation, migration, and metastasis [52]. Two other hypomethylated CpG-linked genes, *VRK2* and *MGAT5B*, have been associated with HCC metastasis [53, 54]. *KCNMA1* has been found to be downregulated in HCC, and its upregulation enhances HCC cell lines’ responsiveness to treatment with sorafenib [55]. Further, *OSBPL5* is reported to be downregulated in HCC [56]. While down regulation of *PAWR* has been found to induce bladder cancer, its upregulation with self-amplifying RNA (saRNA) inhibits cancer cell proliferation by inducing apoptosis [57]. The potential impact of the other genes listed in **Table 2** has not been studied extensively and therefore requires further investigation.

Although HCC is typically diagnosed based on clinical, imaging and/or pathological features, in the present study, we did not aim to establish a diagnostic criterion for metabolic HCC, but rather identify DNA methylation markers that can robustly discriminate metabolic HCC from benign metabolic liver disease. Our aim is that these markers could be combined with clinical biomarkers in future studies to improve diagnosis of HCC in patients with chronic metabolic perturbations, including improving diagnosis in patients with asymptomatic disease. Identifying DNA methylation markers that can discriminate between cancer and non-cancer samples is an important first step in cancer detection in high-risk patients [20]. However, whether the markers identified here are aberrantly methylated in the precancer stage or early cancer development stage of the multistage hepatic tumorigenesis would need to be investigated further before the establishment of a specific criterion for metabolic HCC detection. The identified markers have prospects for clinical translation if confirmed in prospective studies with long-term follow-up and with evaluation of early-stage HCC in the background of metabolic liver disease. Since the methylation markers could be targeted with pyrosequencing or high-performance liquid chromatography, both of which can be done in a clinical laboratory, we expect that their clinical application would be feasible.

Our study has several strengths and limitations. Strengths of the study includes the focus on patients with metabolic liver disease with well-characterized samples sourced through our multicenter international collaboration. We used the 850k EPIC array for screening of differentially methylated CpG positions across the genome, as opposed to the smaller 450k array with limited CpG coverage or targeted assay panels that have been used in prior studies [16, 17, 20, 21, 31, 58]. Our sample size was sufficiently large and enabled separate training and independent validation analyses. To ensure rigor and reduce redundancy (multicollinearity) in CpG selection, we employed LASSO regression analysis with 10-fold cross validation in the training models, which adds to the study’s strengths. Our validation analysis also shows robustness of the models and supports a role of the identified markers in metabolic hepatotumorigenesis. Additionally, we built a separate model focused on only hypermethylated CpGs, which has been done in some studies, but our primary focus was on the combined effect of both the hypermethylated and hypomethylated CpGs. Limitations include our use of leukocyte DNA samples instead of plasma-derived cell-free DNA (cfDNA), for the methylation assay. While we did not have sufficient plasma volume on our patients for the cfDNA assay, we ameliorated this challenge by estimating leukocyte cell type proportions in each participant sample and adjusted for significant cell types in the model used for selecting differentially methylated CpGs. We also did not have data on cirrhosis status or tumor stage, and we could not assess these in the study. The cross-sectional nature of our data cannot preclude reverse causality of the association where the presence of a tumor could alter methylation status. However, such alterations could be useful for early HCC detection if confirmed in longitudinal studies. Further, our study sample is predominantly non-Hispanic White, thus, follow-up studies in a more diverse patient population, preferably using cfDNA and including data on cirrhosis and tumor stage and with larger patient samples would be an improvement.

In summary, we performed an unbiased epigenome-wide screening of differentially methylation markers using germline leukocyte DNA and identified a promising set of CpGs that can discriminate patients with metabolic HCC from cancer-free patients with metabolic liver disease. These markers could aid in HCC surveillance in patients with metabolic perturbations. Although further work is needed to confirm the markers identified here, they could serve as components of an integrative panel that could ultimately improve outcomes for patients with this frequently deadly cance.

## Data Availability

The underlying data used for this analysis has been deposited in the National Center for Biotechnology Information/ Gene Expression Omnibus (GEO) database: GSE281691.

https://www.ncbi.nlm.nih.gov/geo/query/acc.cgi?acc=GSE281691

## Acknowledgements

We thank the patients and controls who made this study possible by providing blood samples and completing the risk factor questionnaires. We also thank the study coordinators and registry staff of the various institutions that contributed data and samples for the study. The study was supported with funding from the U.S. National Institutes of Health | National Cancer Institute to S.O. Antwi (K01 CA237875; P50 CA210964-02A1CEP), A. Singal (U01 CA271887, U01 CA230694), K. Wangensteen (R37 CA259201), and L.R. Roberts (P50 CA210964). The study was further supported with funding from the Cancer Prevention Research Institute of Texas to A. Singal (RP200554). These sponsors did not have any role in the study design, data collection, analysis, or interpretation of the results.

