## Supplementary Tables 1-5 for "Genome-wide DNA methylation markers associated with metabolic liver cancer"

Supplementary Table 1. Differentially methylation positions identified through epigenome-wide discovery analysis in the training data (n=164 CpGs)

| CpG probe | Chromosomal position[a] | Gene | Strand direction | Relation to Island | Cases: Mean beta value | Controls: Mean beta value | Log-fold change[b] | Raw P-value | Q-Value[c] |
|---|---|---|---|---|---|---|---|---|---|
| cg15837522 | Chr8:117892654 | - | - | OpenSea | 0.556246 | 0.514901074 | 0.036868 | 3.10E-09 | 0.002044141 |
| cg08203231 | Chr5:52385556 | *ITGA2* | + | OpenSea | 0.575761 | 0.53707765 | 0.02628 | 5.91E-09 | 0.002044141 |
| cg17408993 | Chr12:65110360 | *GNS* | + | OpenSea | 0.376479 | 0.352074374 | 0.023179 | 1.84E-08 | 0.003210132 |
| cg25433259 | Chr6:159271784 | *OSTCP1* | + | OpenSea | 0.665419 | 0.63835286 | 0.012262 | 1.86E-08 | 0.003210132 |
| cg19639331 | Chr5:158469130 | *EBF1* | + | OpenSea | 0.393416 | 0.378471662 | 0.021791 | 2.60E-08 | 0.003401872 |
| cg16392442 | Chr12:49595954 | - | - | OpenSea | 0.173533 | 0.156960928 | 0.013945 | 2.95E-08 | 0.003401872 |
| cg06457135 | Chr19:5953406 | *RANBP3* | + | OpenSea | 0.408663 | 0.384367856 | 0.020357 | 5.09E-08 | 0.005024759 |
| cg06946797 | Chr16:11422409 | - | + | OpenSea | 0.160856 | 0.18097384 | -0.01745 | 7.73E-08 | 0.006674643 |
| cg16956116 | Chr6:82456876 | *FAM46A* | + | OpenSea | 0.385287 | 0.370858258 | 0.020638 | 1.13E-07 | 0.007395914 |
| cg12910977 | Chr5:138207492 | *LRRTM2;CTNNA1* | - | OpenSea | 0.561393 | 0.517909498 | 0.038322 | 1.18E-07 | 0.007395914 |
| cg19078289 | Chr3:152017293 | *MBNL1* | - | OpenSea | 0.247146 | 0.221092266 | 0.020002 | 1.26E-07 | 0.007395914 |
| cg09605623 | Chr10:112477603 | *RBM20* | + | OpenSea | 0.491197 | 0.478291281 | 0.015343 | 1.43E-07 | 0.007395914 |
| cg18425341 | Chr12:68056997 | - | + | OpenSea | 0.497466 | 0.478812755 | 0.020023 | 1.47E-07 | 0.007395914 |
| cg11387709 | Chr16:65636127 | - | - | OpenSea | 0.583586 | 0.548924573 | 0.037896 | 1.50E-07 | 0.007395914 |
| cg20163056 | Chr19:5005982 | *KDM4B* | - | OpenSea | 0.69829 | 0.683943243 | 0.014829 | 1.84E-07 | 0.00789425 |
| cg21668652 | Chr17:55749538 | *MSI2* | - | OpenSea | 0.389578 | 0.360879699 | 0.026383 | 1.94E-07 | 0.00789425 |
| cg02016405 | Chr2:65297792 | *CEP68* | + | OpenSea | 0.413683 | 0.429687301 | -0.01621 | 1.94E-07 | 0.00789425 |
| cg02023138 | Chr12:51701979 | *BIN2* | - | OpenSea | 0.841818 | 0.831965078 | 0.010813 | 2.53E-07 | 0.009729895 |
| cg13702222 | Chr3:152017240 | *MBNL1* | - | OpenSea | 0.326129 | 0.294143615 | 0.023794 | 2.83E-07 | 0.010300058 |
| cg26457483 | Chr1:120256112 | *PHGDH* | + | S_Shore | 0.608458 | 0.586436171 | 0.02565 | 2.98E-07 | 0.010300058 |
| cg07712198 | Chr5:9015582 | - | - | OpenSea | 0.376364 | 0.363507931 | 0.016718 | 3.15E-07 | 0.010305185 |
| cg07631144 | Chr17:46657393 | *MIR10A* | - | N_Shore | 0.293594 | 0.337616323 | -0.03536 | 3.28E-07 | 0.010305185 |
| cg16417977 | Chr1:9744265 | *PIK3CD-AS2* | + | OpenSea | 0.481606 | 0.452313313 | 0.024771 | 4.11E-07 | 0.012316665 |
| cg20137410 | Chr22:29292645 | *ZNRF3* | + | OpenSea | 0.675531 | 0.662209599 | 0.013739 | 4.28E-07 | 0.012316665 |
| cg03685481 | Chr15:51066872 | - | - | OpenSea | 0.173228 | 0.156469609 | 0.013145 | 4.58E-07 | 0.012474048 |
| cg03629335 | Chr4:100126967 | *ADH6* | + | OpenSea | 0.690358 | 0.657914214 | 0.037768 | 4.69E-07 | 0.012474048 |
| cg17687025 | Chr11:64535291 | *SF1* | + | S_Shore | 0.204457 | 0.185255468 | 0.01395 | 5.13E-07 | 0.012605011 |
| cg04232816 | Chr2:96905390 | *LOC285033* | - | OpenSea | 0.616156 | 0.602390754 | 0.013719 | 5.20E-07 | 0.012605011 |
| cg17745097 | Chr1:11863365 | *MTHFR* | + | N_Shelf | 0.083072 | 0.076597688 | 0.008709 | 5.36E-07 | 0.012605011 |
| cg16993186 | Chr10:11224102 | *CELF2* | - | OpenSea | 0.523266 | 0.497917214 | 0.020422 | 5.49E-07 | 0.012605011 |
| cg09302321 | Chr7:141676574 | - | + | OpenSea | 0.423086 | 0.401807225 | 0.019022 | 5.85E-07 | 0.012605011 |
| cg17938245 | Chr12:80067352 | *PAWR* | - | OpenSea | 0.686657 | 0.693617997 | -0.01191 | 5.92E-07 | 0.012605011 |
| cg05613718 | Chr7:38355100 | - | + | S_Shelf | 0.245969 | 0.221887116 | 0.020049 | 6.02E-07 | 0.012605011 |

Supplementary Table 1 (continued).

| CpG probe | Chromosomal position[a] | Gene | Strand direction | Relation to Island | Cases: Mean beta value | Controls: Mean beta value | Log-fold change[b] | Raw P-value | Q-Value[c] |
|---|---|---|---|---|---|---|---|---|---|
| cg25124943 | Chr10:4117248 | - | + | OpenSea | 0.641321 | 0.64439514 | -0.01111 | 6.44E-07 | 0.013086802 |
| cg19420069 | Chr16:65635834 | - | + | OpenSea | 0.484117 | 0.455348858 | 0.032551 | 7.39E-07 | 0.014600917 |
| cg20911897 | Chr4:37953726 | *TBC1D1* | + | OpenSea | 0.468677 | 0.452628847 | 0.021575 | 7.96E-07 | 0.015021512 |
| cg23712251 | Chr22:36735642 | *MYH9* | - | OpenSea | 0.606072 | 0.57746622 | 0.01247 | 8.10E-07 | 0.015021512 |
| cg26305504 | Chr19:947612 | *ARID3A* | - | Island | 0.487067 | 0.516411296 | -0.01837 | 8.26E-07 | 0.015021512 |
| cg02881681 | Chr17:2099187 | *SMG6* | - | OpenSea | 0.651066 | 0.625052823 | 0.012906 | 9.47E-07 | 0.016784733 |
| cg03746015 | Chr16:11141000 | *CLEC16A* | + | OpenSea | 0.602517 | 0.647432381 | -0.03523 | 9.78E-07 | 0.016893786 |
| cg09175485 | Chr11:67020555 | *KDM2A* | - | S_Shelf | 0.191424 | 0.184898062 | 0.011781 | 1.04E-06 | 0.017474848 |
| cg06690548 | Chr4:139162808 | *SLC7A11* | - | OpenSea | 0.787899 | 0.761387982 | 0.027486 | 1.07E-06 | 0.017687179 |
| cg02505192 | Chr8:19204342 | *SH2D4A* | - | OpenSea | 0.153352 | 0.146019852 | 0.009532 | 1.11E-06 | 0.017789144 |
| cg16661213 | Chr7:142334093 | - | + | OpenSea | 0.317643 | 0.297612704 | 0.01958 | 1.25E-06 | 0.0185162 |
| cg11303839 | Chr7:75405967 | *CCL26* | + | OpenSea | 0.251321 | 0.275198468 | -0.02912 | 1.36E-06 | 0.020351853 |
| cg09225336 | Chr16:65635750 | - | + | OpenSea | 0.510181 | 0.484717422 | 0.028271 | 1.37E-06 | 0.020351853 |
| cg11322819 | Chr19:13694039 | - | - | OpenSea | 0.078308 | 0.085134367 | -0.00499 | 1.38E-06 | 0.020351853 |
| cg12359298 | Chr20:18735103 | *DTD1* | - | OpenSea | 0.598176 | 0.575178846 | 0.020789 | 1.42E-06 | 0.020446459 |
| cg09417716 | Chr14:101538133 | *MEG9* | - | OpenSea | 0.591817 | 0.599956894 | -0.01216 | 1.53E-06 | 0.021145471 |
| cg17894064 | Chr5:112657195 | *MCC* | + | OpenSea | 0.732116 | 0.756946296 | -0.02582 | 1.54E-06 | 0.021145471 |
| cg26491604 | Chr17:46656864 | *HOXB4* | - | S_Shore | 0.27196 | 0.291695616 | -0.01232 | 1.56E-06 | 0.021145471 |
| cg12640435 | Chr1:173031587 | - | + | OpenSea | 0.487494 | 0.518743965 | -0.03346 | 1.60E-06 | 0.021256506 |
| cg10464496 | Chr12:19587815 | - | - | OpenSea | 0.418747 | 0.39978956 | 0.01841 | 1.72E-06 | 0.022425792 |
| cg23569632 | Chr6:44535269 | - | - | OpenSea | 0.396483 | 0.392333144 | 0.011753 | 1.78E-06 | 0.022479767 |
| cg05142445 | Chr14:76309157 | *TTLL5* | + | OpenSea | 0.437191 | 0.428885557 | 0.015559 | 1.83E-06 | 0.022479767 |
| cg22140708 | Chr6:10081664 | - | - | OpenSea | 0.782068 | 0.771703978 | 0.015495 | 1.83E-06 | 0.022479767 |
| cg21650861 | Chr3:127299468 | *TPRA1* | - | OpenSea | 0.499458 | 0.492590304 | 0.009068 | 1.85E-06 | 0.022479767 |
| cg20989855 | Chr4:20985927 | *KCNIP4* | - | OpenSea | 0.220962 | 0.204735145 | 0.016429 | 1.90E-06 | 0.022643892 |
| cg08311647 | Chr10:11596182 | *USP6NL* | + | OpenSea | 0.320349 | 0.33414088 | -0.0155 | 2.03E-06 | 0.022687772 |
| cg03096649 | Chr7:130056977 | *CEP41* | - | OpenSea | 0.801817 | 0.784614838 | 0.011534 | 2.03E-06 | 0.022687772 |
| cg10946755 | Chr7:36764613 | *AOAH* | + | OpenSea | 0.473558 | 0.455254575 | 0.016846 | 2.11E-06 | 0.022687772 |
| cg09858955 | Chr2:58135951 | *VRK2* | + | OpenSea | 0.357966 | 0.397888502 | -0.03021 | 2.11E-06 | 0.022687772 |
| cg21618197 | Chr9:35611399 | *CD72* | + | OpenSea | 0.148248 | 0.137455566 | 0.009227 | 2.20E-06 | 0.022687772 |
| cg22051853 | Chr7:2647589 | *IQCE* | + | S_Shore | 0.370412 | 0.393579065 | -0.01201 | 2.22E-06 | 0.022687772 |
| cg00506299 | Chr3:16469127 | *RFTN1* | - | OpenSea | 0.573299 | 0.608105769 | -0.02132 | 2.23E-06 | 0.022687772 |
| cg05903720 | Chr14:104663241 | - | - | OpenSea | 0.671042 | 0.656044052 | 0.018513 | 2.23E-06 | 0.022687772 |

**Supplementary Table 1** (continued).

| CpG probe | Chromosomal position[a] | Gene | Strand direction | Relation to Island | Cases: Mean beta value | Controls: Mean beta value | Log-fold change[b] | Raw P-value | Q-Value[c] |
|---|---|---|---|---|---|---|---|---|---|
| cg20150812 | Chr9:124029753 | *GSN* | + | OpenSea | 0.631561 | 0.619989602 | 0.014187 | 2.25E-06 | 0.022687772 |
| cg27060295 | Chr7:8171773 | *ICA1* | - | OpenSea | 0.589007 | 0.566206056 | 0.009945 | 2.26E-06 | 0.022687772 |
| cg17602903 | Chr1:27986074 | - | + | N_Shore | 0.630259 | 0.615597561 | 0.010167 | 2.28E-06 | 0.022687772 |
| cg24356337 | Chr5:93758545 | *KIAA0825* | - | OpenSea | 0.534072 | 0.568868039 | -0.02098 | 2.30E-06 | 0.022687772 |
| cg25072592 | Chr14:75355586 | *DLST* | - | OpenSea | 0.531999 | 0.512581172 | 0.021077 | 2.34E-06 | 0.022776254 |
| cg18265326 | Chr16:65635738 | - | - | OpenSea | 0.517625 | 0.494709201 | 0.026139 | 2.53E-06 | 0.02431963 |
| cg02536065 | Chr11:128616196 | *FLI1* | + | OpenSea | 0.66211 | 0.652932542 | 0.014065 | 2.64E-06 | 0.024712463 |
| cg08972170 | Chr7:30185776 | *C7orf41* | - | OpenSea | 0.519695 | 0.496380982 | 0.024392 | 2.65E-06 | 0.024712463 |
| cg17702370 | Chr17:79283128 | *C17orf55* | + | N_Shore | 0.157749 | 0.148590665 | 0.011935 | 2.79E-06 | 0.025684613 |
| cg13095216 | Chr22:27044694 | - | + | OpenSea | 0.409323 | 0.404558605 | 0.011846 | 2.83E-06 | 0.025770792 |
| cg26129664 | Chr10:134232399 | - | + | OpenSea | 0.398212 | 0.388355911 | 0.014862 | 2.93E-06 | 0.026269956 |
| cg04509882 | Chr3:184038317 | *EIF4G1* | - | OpenSea | 0.6622 | 0.654542669 | 0.016428 | 2.99E-06 | 0.026495592 |
| cg21946374 | Chr5:1108401 | *SLC12A7* | - | N_Shore | 0.310985 | 0.332704302 | -0.00937 | 3.14E-06 | 0.027304054 |
| cg26532527 | Chr13:100033381 | *UBAC2;MIR548AN* | - | OpenSea | 0.611341 | 0.584185807 | 0.010693 | 3.16E-06 | 0.027304054 |
| cg18512769 | Chr22:50825141 | *PPP6R2* | + | OpenSea | 0.511008 | 0.490269822 | 0.018416 | 3.25E-06 | 0.027383603 |
| cg03337382 | Chr10:125960385 | - | + | OpenSea | 0.691471 | 0.711367438 | -0.01304 | 3.28E-06 | 0.027383603 |
| cg02447854 | Chr11:72498689 | *STARD10* | + | OpenSea | 0.327628 | 0.351082653 | -0.0116 | 3.29E-06 | 0.027383603 |
| cg17782974 | Chr10:104406990 | *TRIM8* | - | S_Shelf | 0.335922 | 0.31012413 | 0.019549 | 3.42E-06 | 0.028138379 |
| cg07209338 | Chr21:39844537 | *ERG* | + | OpenSea | 0.553904 | 0.545766172 | 0.015352 | 3.47E-06 | 0.02824466 |
| cg17320738 | Chr8:32726179 | - | + | OpenSea | 0.128313 | 0.123101263 | 0.007234 | 3.56E-06 | 0.028266416 |
| cg26715529 | Chr13:108877982 | *ABHD13* | + | OpenSea | 0.073483 | 0.069334155 | 0.005307 | 3.56E-06 | 0.028266416 |
| cg20218040 | Chr6:14369697 | - | - | OpenSea | 0.784529 | 0.761995514 | 0.016775 | 3.60E-06 | 0.028266416 |
| cg09755313 | Chr12:56532555 | *ESYT1* | - | OpenSea | 0.762447 | 0.743037167 | 0.014373 | 3.67E-06 | 0.028491454 |
| cg06245989 | Chr11:16828408 | *PLEKHA7* | + | OpenSea | 0.519757 | 0.509441661 | 0.013968 | 3.75E-06 | 0.02878925 |
| cg12600265 | Chr7:5422883 | *TNRC18* | - | OpenSea | 0.579376 | 0.56715465 | 0.016392 | 3.88E-06 | 0.029077501 |
| cg26403045 | Chr16:2576370 | *AMDHD2* | - | N_Shore | 0.451909 | 0.444991602 | 0.01064 | 3.91E-06 | 0.029077501 |
| cg10934068 | Chr6:509762 | *EXOC2* | + | OpenSea | 0.585621 | 0.593797126 | -0.0261 | 3.92E-06 | 0.029077501 |
| cg11219937 | Chr12:64664370 | *C12orf56* | - | OpenSea | 0.649999 | 0.63470156 | 0.020001 | 4.02E-06 | 0.029077501 |
| cg20294744 | Chr13:48978285 | *RB1* | - | OpenSea | 0.723948 | 0.759613269 | -0.03002 | 4.02E-06 | 0.029077501 |
| cg26669602 | Chr20:62243857 | *GMEB2* | + | OpenSea | 0.353152 | 0.375479285 | -0.00902 | 4.04E-06 | 0.029077501 |
| cg11096441 | Chr20:49548515 | *ADNP* | + | Island | 0.106509 | 0.097115099 | 0.008398 | 4.22E-06 | 0.030052748 |
| cg13039251 | Chr5:32018601 | *PDZD2* | - | OpenSea | 0.585972 | 0.558399722 | 0.033387 | 4.34E-06 | 0.030616329 |
| cg01148781 | Chr8:99963021 | *OSR2* | + | S_Shore | 0.127759 | 0.148460038 | -0.02274 | 4.70E-06 | 0.032620454 |

Supplementary Table 1 (continued).

| CpG probe | Chromosomal position[a] | Gene | Strand direction | Relation to Island | Cases: Mean beta values | Controls: Mean beta value | Log-fold change[b] | Raw P-value | Q-Value[c] |
|---|---|---|---|---|---|---|---|---|---|
| cg08483768 | Chr16:86304619 | - | + | OpenSea | 0.509459 | 0.482372721 | 0.034481 | 4.72E-06 | 0.032620454 |
| cg01072106 | Chr9:138952311 | *NACC2* | - | OpenSea | 0.669974 | 0.690908509 | -0.01338 | 4.89E-06 | 0.032907508 |
| cg00941658 | Chr20:58563977 | *CDH26* | - | OpenSea | 0.454676 | 0.449741906 | 0.011979 | 4.92E-06 | 0.032907508 |
| cg15334909 | Chr2:145253398 | *ZEB2* | + | OpenSea | 0.350356 | 0.342368411 | 0.015226 | 4.95E-06 | 0.032907508 |
| cg09316997 | Chr14:91874913 | *CCDC88C* | - | OpenSea | 0.588556 | 0.563127383 | 0.011506 | 5.05E-06 | 0.032907508 |
| cg02650908 | Chr17:74889830 | *MGAT5B* | + | OpenSea | 0.716946 | 0.738717762 | -0.02188 | 5.07E-06 | 0.032907508 |
| cg25289880 | Chr1:13856326 | - | + | OpenSea | 0.117508 | 0.106232905 | 0.011847 | 5.07E-06 | 0.032907508 |
| cg08760843 | Chr8:28196932 | *PNOC* | - | OpenSea | 0.338776 | 0.360968378 | -0.01226 | 5.09E-06 | 0.032907508 |
| cg01592226 | Chr22:36740752 | *MYH9* | + | OpenSea | 0.70565 | 0.678761893 | 0.011437 | 5.23E-06 | 0.033479738 |
| cg08834436 | Chr22:27831818 | - | + | N_Shelf | 0.330845 | 0.366664689 | -0.03701 | 5.43E-06 | 0.034231142 |
| cg04643650 | Chr12:7008890 | - | + | OpenSea | 0.461004 | 0.445746492 | 0.022339 | 5.47E-06 | 0.034231142 |
| cg08343240 | Chr5:158886936 | *LOC285627* | - | OpenSea | 0.655232 | 0.670316745 | -0.02149 | 5.50E-06 | 0.034231142 |
| cg18765390 | Chr8:49529851 | *LOC101929268* | + | OpenSea | 0.253121 | 0.243296848 | 0.014894 | 5.67E-06 | 0.034976695 |
| cg10636929 | Chr8:134595388 | - | - | OpenSea | 0.80611 | 0.785885147 | 0.012035 | 5.84E-06 | 0.035030841 |
| cg23206630 | Chr15:62942319 | *TLN2* | - | OpenSea | 0.50899 | 0.494553951 | 0.02363 | 5.84E-06 | 0.035030841 |
| cg03593369 | Chr2:43456557 | - | - | S_Shore | 0.293231 | 0.273533246 | 0.013588 | 5.89E-06 | 0.035030841 |
| cg13975855 | Chr17:46652550 | *HOXB3* | - | N_Shore | 0.761648 | 0.783370366 | -0.0168 | 5.93E-06 | 0.035030841 |
| cg21953769 | Chr19:30168190 | - | - | S_Shelf | 0.528807 | 0.516187977 | 0.006921 | 5.93E-06 | 0.035030841 |
| cg06548416 | Chr1:24438703 | *MYOM3* | - | OpenSea | 0.232424 | 0.205557918 | 0.028631 | 6.24E-06 | 0.036283587 |
| cg23029198 | Chr2:146510791 | - | + | OpenSea | 0.79024 | 0.795595849 | -0.00816 | 6.25E-06 | 0.036283587 |
| cg02862897 | Chr17:79793088 | *DYSFIP1* | + | S_Shore | 0.634194 | 0.651334328 | -0.01362 | 6.30E-06 | 0.036283587 |
| cg18591228 | Chr11:3175552 | *OSBPL5* | - | OpenSea | 0.35349 | 0.374484016 | -0.00972 | 6.57E-06 | 0.037504117 |
| cg06198776 | Chr13:73557424 | *PIBF1* | + | OpenSea | 0.870813 | 0.881560245 | -0.00838 | 6.73E-06 | 0.037855611 |
| cg03442235 | Chr6:15369118 | *JARID2* | - | OpenSea | 0.721698 | 0.709886922 | 0.016373 | 6.75E-06 | 0.037855611 |
| cg26072749 | Chr17:46657274 | *MIR10A* | + | N_Shore | 0.157821 | 0.176169992 | -0.01432 | 6.87E-06 | 0.037855611 |
| cg11994115 | Chr19:40360856 | *FCGBP* | + | N_Shore | 0.643067 | 0.618532712 | 0.02645 | 6.87E-06 | 0.037855611 |
| cg15081698 | Chr6:101847050 | *GRIK2* | - | Island | 0.123313 | 0.111708053 | 0.010803 | 6.97E-06 | 0.037855611 |
| cg18496725 | Chr4:68788615 | *TMPRSS11A* | - | OpenSea | 0.751069 | 0.776024586 | -0.0244 | 6.97E-06 | 0.037855611 |
| cg08553572 | Chr19:947765 | *ARID3A* | - | Island | 0.312474 | 0.339034001 | -0.01463 | 7.01E-06 | 0.037855611 |
| cg08146029 | Chr5:130868107 | *RAPGEF6* | + | OpenSea | 0.481138 | 0.493976427 | -0.02088 | 7.15E-06 | 0.038297218 |
| cg05014727 | Chr10:6214016 | *PFKFB3* | + | OpenSea | 0.286269 | 0.317048077 | -0.01989 | 7.25E-06 | 0.038525675 |
| cg23852535 | Chr17:8857258 | *PIK3R5* | + | OpenSea | 0.837875 | 0.83078653 | 0.00738 | 7.32E-06 | 0.038533382 |
| cg14217303 | Chr15:85177537 | *SCAND2P* | + | S_Shore | 0.25646 | 0.247047364 | 0.009832 | 7.36E-06 | 0.038533382 |

Supplementary Table 1 (continued).

| CpG Probe | Chromosomal position[a] | Gene | Strand direction | Relation to Island | Cases: Mean beta value | Controls: Mean beta value | Log-fold change[b] | Raw P-value | Q-Value[c] |
|---|---|---|---|---|---|---|---|---|---|
| cg12256648 | Chr3:143752097 | - | - | OpenSea | 0.420996 | 0.44279351 | -0.02348 | 7.53E-06 | 0.038928697 |
| cg08841898 | Chr12:27717865 | *PPFIBP1* | + | OpenSea | 0.687452 | 0.700223035 | -0.00927 | 7.55E-06 | 0.038928697 |
| cg00418799 | Chr6:169989419 | *WDR27* | - | OpenSea | 0.354678 | 0.319531745 | 0.036853 | 7.63E-06 | 0.039052566 |
| cg16615151 | Chr3:111409324 | *PLCXD2* | - | OpenSea | 0.515153 | 0.49645295 | 0.017994 | 7.70E-06 | 0.03911141 |
| cg10341940 | Chr18:76822780 | - | + | OpenSea | 0.823907 | 0.829772334 | -0.00657 | 7.77E-06 | 0.039223939 |
| cg08922729 | Chr1:21913557 | - | - | OpenSea | 0.427264 | 0.450398615 | -0.01684 | 7.97E-06 | 0.039931646 |
| cg22111527 | Chr11:69260136 | - | - | S_Shore | 0.346745 | 0.372537528 | -0.01231 | 8.03E-06 | 0.039931646 |
| cg05083539 | Chr2:219150776 | *PNKD;TMBIM1* | + | OpenSea | 0.392891 | 0.380528504 | 0.010155 | 8.42E-06 | 0.041571359 |
| cg14510926 | Chr2:174877357 | - | - | N_Shore | 0.440952 | 0.42779554 | 0.018399 | 8.51E-06 | 0.041688516 |
| cg25153204 | Chr10:79291246 | *KCNMA1* | + | OpenSea | 0.849993 | 0.858352427 | -0.01105 | 8.58E-06 | 0.041688516 |
| cg09792879 | Chr2:202126492 | *CASP8* | + | OpenSea | 0.195114 | 0.181069997 | 0.012961 | 8.62E-06 | 0.041688516 |
| cg03282313 | Chr2:113545053 | - | + | OpenSea | 0.396612 | 0.41773664 | -0.00945 | 8.90E-06 | 0.042680011 |
| cg03366951 | Chr3:39302545 | - | + | OpenSea | 0.163064 | 0.153998609 | 0.01408 | 8.95E-06 | 0.042680011 |
| cg11123646 | Chr13:99215537 | *STK24* | - | OpenSea | 0.538028 | 0.540752339 | -0.01194 | 9.04E-06 | 0.042790928 |
| cg08373547 | Chr14:52794541 | *PTGER2* | - | OpenSea | 0.233357 | 0.25315212 | -0.02029 | 9.11E-06 | 0.042834525 |
| cg11047325 | Chr17:76354934 | *SOCS3* | - | Island | 0.469419 | 0.51306814 | -0.03752 | 9.17E-06 | 0.042834525 |
| cg14025021 | Chr15:43809510 | *MAP1A* | + | OpenSea | 0.518398 | 0.507830612 | 0.015233 | 9.34E-06 | 0.043320763 |
| cg17475445 | Chr22:31344026 | *MORC2* | - | OpenSea | 0.604246 | 0.583989987 | 0.011105 | 9.51E-06 | 0.043827535 |
| cg21668714 | Chr16:309450 | *ITFG3* | - | OpenSea | 0.514544 | 0.542202056 | -0.02349 | 1.01E-05 | 0.046056662 |
| cg04953230 | Chr11:65273641 | *MALAT1* | - | OpenSea | 0.546102 | 0.521952988 | 0.018645 | 1.01E-05 | 0.046056662 |
| cg03741619 | Chr17:3438918 | *TRPV3* | - | Island | 0.07389 | 0.068182814 | 0.005446 | 1.03E-05 | 0.046105568 |
| cg03967504 | Chr2:219248734 | *SLC11A1* | + | N_Shelf | 0.426362 | 0.442762329 | -0.00939 | 1.03E-05 | 0.046105568 |
| cg01601658 | Chr6:168785524 | - | - | OpenSea | 0.307781 | 0.288746997 | 0.019683 | 1.04E-05 | 0.046456723 |
| cg06088069 | Chr14:75895604 | *JDP2* | + | S_Shore | 0.4506 | 0.441855125 | 0.012009 | 1.07E-05 | 0.047404809 |
| cg20397849 | Chr11:67051709 | *ADRBK1* | - | N_Shore | 0.35185 | 0.374128739 | -0.00865 | 1.09E-05 | 0.048202286 |
| cg08635097 | Chr13:44833857 | - | + | OpenSea | 0.129426 | 0.123140906 | 0.007551 | 1.11E-05 | 0.048688472 |
| cg21539223 | Chr5:112312093 | *DCP2* | - | N_Shore | 0.049279 | 0.044646758 | 0.004761 | 1.13E-05 | 0.048994621 |
| cg00867472 | Chr1:156714808 | *HDGF* | - | S_Shelf | 0.587221 | 0.578165548 | 0.01162 | 1.15E-05 | 0.048994621 |
| cg08799032 | Chr2:46229638 | *PRKCE* | - | OpenSea | 0.519696 | 0.503732755 | 0.016215 | 1.15E-05 | 0.048994621 |
| cg08960830 | Chr11:75047180 | *ARRB1* | - | OpenSea | 0.699905 | 0.692487485 | 0.00905 | 1.16E-05 | 0.048994621 |
| cg14476101 | Chr1:120255992 | *PHGDH* | - | S_Shore | 0.557482 | 0.537611327 | 0.023292 | 1.16E-05 | 0.048994621 |
| cg26221987 | Chr10:4289330 | - | + | OpenSea | 0.315829 | 0.329928139 | -0.01545 | 1.16E-05 | 0.048994621 |

[a]Positions are based on the human reference genome assembly GRCh38.
[b]Fold change comparing beta estimates between cases and controls.
[c]False-discovery rate (FDR)-adjusted p-value.

**Supplementary Table 2**: CpGs used for the elaborate multifactorial model in Figure 3C (n=44 CpGs).

| CpG probe | Chromosomal position[a] | Gene | Relation to Island | Cases: Mean beta value | Controls: Mean beta value | Log-fold change[b] | Raw P-value | Q-Value[c] |
|---|---|---|---|---|---|---|---|---|
| cg25433259 | Chr6:159271784 | *OSTCP1* | OpenSea | 0.66541858 | 0.63835286 | 0.01226212 | 1.86E-08 | 0.00321013 |
| cg16392442 | Chr12:49595954 | - | OpenSea | 0.17353326 | 0.15696093 | 0.01394544 | 2.95E-08 | 0.00340187 |
| cg06457135 | Chr19:5953406 | *RANBP3* | OpenSea | 0.40866314 | 0.38436786 | 0.02035666 | 5.09E-08 | 0.00502476 |
| cg02023138 | Chr12:51701979 | *BIN2* | OpenSea | 0.84181837 | 0.83196508 | 0.01081285 | 2.53E-07 | 0.00972989 |
| cg07631144 | Chr17:46657393 | *MIR10A* | N_Shore | 0.29359393 | 0.33761632 | -0.0353619 | 3.28E-07 | 0.01030519 |
| cg03629335 | Chr4:100126967 | *LOC100507053;ADH6* | OpenSea | 0.69035821 | 0.65791421 | 0.03776824 | 4.69E-07 | 0.01247405 |
| cg17745097 | Chr1:11863365 | *MTHFR* | N_Shelf | 0.08307216 | 0.07659769 | 0.00870919 | 5.36E-07 | 0.01260501 |
| cg17938245 | Chr12:80067352 | *PAWR* | OpenSea | 0.68665745 | 0.693618 | -0.0119055 | 5.92E-07 | 0.01260501 |
| cg20911897 | Chr4:37953726 | *TBC1D1* | OpenSea | 0.46867666 | 0.45262885 | 0.02157469 | 7.96E-07 | 0.01502151 |
| cg11322819 | Chr19:13694039 | - | OpenSea | 0.07830778 | 0.08513437 | -0.0049903 | 1.38E-06 | 0.02035185 |
| cg17894064 | Chr5:112657195 | *MCC* | OpenSea | 0.73211569 | 0.7569463 | -0.0258219 | 1.54E-06 | 0.02114547 |
| cg22140708 | Chr6:10081664 | - | OpenSea | 0.78206787 | 0.77170398 | 0.0154948 | 1.83E-06 | 0.02247977 |
| cg20989855 | Chr4:20985927 | *KCNIP4* | OpenSea | 0.22096169 | 0.20473515 | 0.0164293 | 1.90E-06 | 0.02264389 |
| cg08311647 | Chr10:11596182 | *USP6NL* | OpenSea | 0.32034876 | 0.33414088 | -0.0155017 | 2.03E-06 | 0.02268777 |
| cg03096649 | Chr7:130056977 | *CEP41* | OpenSea | 0.80181661 | 0.78461484 | 0.01153378 | 2.03E-06 | 0.02268777 |
| cg09858955 | Chr2:58135951 | *VRK2* | OpenSea | 0.35796606 | 0.3978885 | -0.0302066 | 2.11E-06 | 0.02268777 |
| cg20150812 | Chr9:124029753 | *GSN* | OpenSea | 0.63156105 | 0.6199896 | 0.01418724 | 2.25E-06 | 0.02268777 |
| cg25072592 | Chr14:75355586 | *DLST* | OpenSea | 0.53199877 | 0.51258117 | 0.0210767 | 2.34E-06 | 0.02277625 |
| cg17702370 | Chr17:79283128 | *C17orf55* | N_Shore | 0.15774941 | 0.14859067 | 0.0119348 | 2.79E-06 | 0.02568461 |
| cg10934068 | Chr6:509762 | *EXOC2* | OpenSea | 0.58562148 | 0.59379713 | -0.0261018 | 3.92E-06 | 0.0290775 |
| cg01148781 | Chr8: 99963021 | *OSR2* | S_Shore | 0.127759 | 0.14846004 | -0.0227403 | 4.70E-06 | 0.03262045 |
| cg08483768 | Chr16:86304619 | - | OpenSea | 0.50945873 | 0.48237272 | 0.0344807 | 4.72E-06 | 0.03262045 |
| cg01072106 | Chr9:138952311 | *NACC2* | OpenSea | 0.66997388 | 0.69090851 | -0.0133822 | 4.89E-06 | 0.03290751 |
| cg09316997 | Chr14:91874913 | *CCDC88C* | OpenSea | 0.58855649 | 0.56312738 | 0.01150639 | 5.05E-06 | 0.03290751 |
| cg08834436 | Chr22:27831818 | - | N_Shelf | 0.33084528 | 0.36666469 | -0.0370071 | 5.43E-06 | 0.03423114 |
| cg13975855 | Chr17:46652550 | *HOXB3* | N_Shore | 0.7616477 | 0.78337037 | -0.0167974 | 5.93E-06 | 0.03503084 |
| cg06548416 | Chr1:24438703 | *MYOM3* | OpenSea | 0.23242444 | 0.20555792 | 0.02863055 | 6.24E-06 | 0.03628359 |
| cg23029198 | Chr2:146510791 | - | OpenSea | 0.79023973 | 0.79559585 | -0.0081623 | 6.25E-06 | 0.03628359 |
| cg18591228 | Chr11:3175552 | *OSBPL5* | OpenSea | 0.35349018 | 0.37448402 | -0.0097212 | 6.57E-06 | 0.03750412 |
| cg06198776 | Chr13:73557424 | *PIBF1* | OpenSea | 0.87081259 | 0.88156024 | -0.008382 | 6.73E-06 | 0.03785561 |
| cg26072749 | Chr17:46657274 | *MIR10A* | N_Shore | 0.15782119 | 0.17616999 | -0.0143176 | 6.87E-06 | 0.03785561 |
| cg11994115 | Chr19:40360856 | *FCGBP* | N_Shore | 0.64306667 | 0.61853271 | 0.02644958 | 6.87E-06 | 0.03785561 |
| cg15081698 | Chr6:101847050 | *GRIK2* | Island | 0.12331257 | 0.11170805 | 0.01080277 | 6.97E-06 | 0.03785561 |

Supplementary Table 2 (continued).

| CpG probe | Chromosomal position[a] | Gene | Relation to Island | Cases: Mean beta value | Controls: Mean beta value | Log-fold change[b] | Raw P-value | Q-Value[c] |
|---|---|---|---|---|---|---|---|---|
| cg18496725 | Chr4:68788615 | *TMPRSS11A* | OpenSea | 0.75106914 | 0.77602459 | -0.0244003 | 6.97E-06 | 0.03785561 |
| cg23852535 | Chr17:8857258 | *PIK3R5* | OpenSea | 0.83787502 | 0.83078653 | 0.00737971 | 7.32E-06 | 0.03853338 |
| cg12256648 | Chr3:143752097 | - | OpenSea | 0.42099641 | 0.44279351 | -0.0234836 | 7.53E-06 | 0.0389287 |
| cg08841898 | Chr12:27717865 | *PPFIBP1* | OpenSea | 0.68745191 | 0.70022304 | -0.0092687 | 7.55E-06 | 0.0389287 |
| cg10341940 | Chr18:76822780 | - | OpenSea | 0.82390723 | 0.82977233 | -0.006565 | 7.77E-06 | 0.03922394 |
| cg03366951 | Chr3:39302545 | - | OpenSea | 0.16306437 | 0.15399861 | 0.01408014 | 8.95E-06 | 0.04268001 |
| cg03741619 | Chr17:3438918 | *TRPV3* | Island | 0.07389002 | 0.06818281 | 0.0054458 | 1.03E-05 | 0.04610557 |
| cg01601658 | Chr6:168785524 | - | OpenSea | 0.30778145 | 0.288747 | 0.01968328 | 1.04E-05 | 0.04645672 |
| cg08635097 | Chr13:44833857 | - | OpenSea | 0.12942612 | 0.12314091 | 0.00755129 | 1.11E-05 | 0.04868847 |
| cg21539223 | Chr5: 112312093 | *DCP2* | N_Shore | 0.04927896 | 0.04464676 | 0.00476079 | 1.13E-05 | 0.04899462 |
| cg08960830 | Chr11:75047180 | *ARRB1;MIR326* | OpenSea | 0.69990481 | 0.69248749 | 0.00905002 | 1.16E-05 | 0.04899462 |

[a]Positions are based on the human reference genome assembly GRCh38.
[b]Fold change comparing beta estimates between cases and controls.
[c]False-discovery rate (FDR)-adjusted p-value.

**Supplementary Table 3.** Hypermethylated CpGs selected for the CpG only model (n=42 CpGs)

| CpG Probe | Chromosomal position[a] | Gene | Relation to Island | Cases: Mean beta value | Controls: Mean beta value | Log-fold change[b] | Raw P-value | Q-Value[c] |
|---|---|---|---|---|---|---|---|---|
| cg15837522 | Chr8:117892654 | - | OpenSea | 0.55624591 | 0.51490107 | 0.03686814 | 3.10E-09 | 0.00204414 |
| cg25433259 | Chr6:159271784 | *OSTCP1* | OpenSea | 0.66541858 | 0.63835286 | 0.01226212 | 1.86E-08 | 0.00321013 |
| cg16392442 | Chr12:49595954 | - | OpenSea | 0.17353326 | 0.15696093 | 0.01394544 | 2.95E-08 | 0.00340187 |
| cg06457135 | Chr19:5953406 | *RANBP3* | OpenSea | 0.40866314 | 0.38436786 | 0.02035666 | 5.09E-08 | 0.00502476 |
| cg12910977 | Chr5:138207492 | *LRRTM2;CTNNA1* | OpenSea | 0.56139306 | 0.5179095 | 0.03832187 | 1.18E-07 | 0.00739591 |
| cg20163056 | Chr19:5005982 | *KDM4B* | OpenSea | 0.69828989 | 0.68394324 | 0.01482862 | 1.84E-07 | 0.00789425 |
| cg02023138 | Chr12:51701979 | *BIN2* | OpenSea | 0.84181837 | 0.83196508 | 0.01081285 | 2.53E-07 | 0.00972989 |
| cg20137410 | Chr22:29292645 | *ZNRF3* | OpenSea | 0.67553079 | 0.6622096 | 0.01373916 | 4.28E-07 | 0.01231666 |
| cg03629335 | Chr4:100126967 | *LOC100507053;ADH6* | OpenSea | 0.69035821 | 0.65791421 | 0.03776824 | 4.69E-07 | 0.01247405 |
| cg17687025 | Chr11:64535291 | *SF1* | S_Shore | 0.20445708 | 0.18525547 | 0.01395003 | 5.13E-07 | 0.01260501 |
| cg17745097 | Chr1:11863365 | *MTHFR* | N_Shelf | 0.08307216 | 0.07659769 | 0.00870919 | 5.36E-07 | 0.01260501 |
| cg09302321 | Chr7:141676574 | - | OpenSea | 0.42308575 | 0.40180723 | 0.01902244 | 5.85E-07 | 0.01260501 |
| cg05613718 | Chr7:38355100 | - | S_Shelf | 0.245969 | 0.22188712 | 0.02004878 | 6.02E-07 | 0.01260501 |
| cg20911897 | Chr4:37953726 | *TBC1D1* | OpenSea | 0.46867666 | 0.45262885 | 0.02157469 | 7.96E-07 | 0.01502151 |
| cg06690548 | Chr4:139162808 | *SLC7A11* | OpenSea | 0.78789942 | 0.76138798 | 0.0274857 | 1.07E-06 | 0.01768718 |
| cg22140708 | Chr6:10081664 | - | OpenSea | 0.78206787 | 0.77170398 | 0.0154948 | 1.83E-06 | 0.02247977 |
| cg20989855 | Chr4:20985927 | *KCNIP4* | OpenSea | 0.22096169 | 0.20473515 | 0.0164293 | 1.90E-06 | 0.02264389 |
| cg03096649 | Chr7:130056977 | *CEP41* | OpenSea | 0.80181661 | 0.78461484 | 0.01153378 | 2.03E-06 | 0.02268777 |
| cg05903720 | Chr14:104663241 | - | OpenSea | 0.67104233 | 0.65604405 | 0.01851333 | 2.23E-06 | 0.02268777 |
| cg20150812 | Chr9:124029753 | *GSN* | OpenSea | 0.63156105 | 0.6199896 | 0.01418724 | 2.25E-06 | 0.02268777 |
| cg17602903 | Chr1:27986074 | - | N_Shore | 0.63025904 | 0.61559756 | 0.01016734 | 2.28E-06 | 0.02268777 |
| cg17702370 | Chr17:79283128 | *C17orf55* | N_Shore | 0.15774941 | 0.14859067 | 0.0119348 | 2.79E-06 | 0.02568461 |
| cg17320738 | Chr8:32726179 | - | OpenSea | 0.12831339 | 0.12310126 | 0.0072337 | 3.56E-06 | 0.02826642 |
| cg20218040 | Chr6:14369697 | - | OpenSea | 0.78452904 | 0.76199551 | 0.01677532 | 3.60E-06 | 0.02826642 |
| cg13039251 | Chr5:32018601 | *PDZD2* | OpenSea | 0.58597178 | 0.55839972 | 0.03338714 | 4.34E-06 | 0.03061633 |
| cg08483768 | Chr16:86304619 | - | OpenSea | 0.50945873 | 0.48237272 | 0.0344807 | 4.72E-06 | 0.03262045 |
| cg09316997 | Chr14:91874913 | *CCDC88C* | OpenSea | 0.58855649 | 0.56312738 | 0.01150639 | 5.05E-06 | 0.03290751 |
| cg10636929 | Chr8:134595388 | - | OpenSea | 0.80611044 | 0.78588515 | 0.01203522 | 5.84E-06 | 0.03503084 |
| cg03593369 | Chr2:43456557 | - | S_Shore | 0.29323094 | 0.27353325 | 0.01358761 | 5.89E-06 | 0.03503084 |
| cg21953769 | Chr19:30168190 | - | S_Shelf | 0.52880666 | 0.51618798 | 0.00692138 | 5.93E-06 | 0.03503084 |
| cg06548416 | Chr1:24438703 | *MYOM3* | OpenSea | 0.23242444 | 0.20555792 | 0.02863055 | 6.24E-06 | 0.03628359 |
| cg03442235 | Chr6:15369118 | *JARID2* | OpenSea | 0.72169752 | 0.70988692 | 0.01637297 | 6.75E-06 | 0.03785561 |
| cg11994115 | Chr19:40360856 | *FCGBP* | N_Shore | 0.64306667 | 0.61853271 | 0.02644958 | 6.87E-06 | 0.03785561 |

**Supplementary Table 3** (continued).

| CpG Probe | Chromosomal position[a] | Gene | Relation to Island | Cases: Mean beta value | Controls: Mean beta value | Log-fold change[b] | Raw P-value | Q-Value[c] |
|---|---|---|---|---|---|---|---|---|
| cg15081698 | Chr6:101847050 | *GRIK2* | Island | 0.12331257 | 0.11170805 | 0.01080277 | 6.97E-06 | 0.03785561 |
| cg23852535 | Chr17:8857258 | *PIK3R5* | OpenSea | 0.83787502 | 0.83078653 | 0.00737971 | 7.32E-06 | 0.03853338 |
| cg14217303 | Chr15:85177537 | *SCAND2P* | S_Shore | 0.25645962 | 0.24704736 | 0.00983248 | 7.36E-06 | 0.03853338 |
| cg03366951 | Chr3: 39302545 | - | OpenSea | 0.16306437 | 0.15399861 | 0.01408014 | 8.95E-06 | 0.04268001 |
| cg03741619 | Chr17:3438918 | *TRPV3* | Island | 0.07389002 | 0.06818281 | 0.0054458 | 1.03E-05 | 0.04610557 |
| cg01601658 | Chr6:168785524 | - | OpenSea | 0.30778145 | 0.288747 | 0.01968328 | 1.04E-05 | 0.04645672 |
| cg08635097 | Chr13:44833857 | - | OpenSea | 0.12942612 | 0.12314091 | 0.00755129 | 1.11E-05 | 0.04868847 |
| cg21539223 | Chr5:112312093 | *DCP2* | N_Shore | 0.04927896 | 0.04464676 | 0.00476079 | 1.13E-05 | 0.04899462 |
| cg08960830 | Chr11:75047180 | *ARRB1;MIR326* | OpenSea | 0.69990481 | 0.69248749 | 0.00905002 | 1.16E-05 | 0.04899462 |

[a]Positions are based on the human reference genome assembly GRCh38.
[b]Fold change comparing beta estimates between cases and controls.
[c]False-discovery rate (FDR)-adjusted p-value.

Supplementary Table 4. Hypermethylated CpGs used for multifactorial modeling together with demographic and clinical data (n=40 CpGs).

| CpG Probe | Chromosomal position[a] | Gene | Relation to Island | Cases: Mean beta value | Controls: Mean beta value | Log-fold change[b] | Raw P-value | Q-Value[c] |
|---|---|---|---|---|---|---|---|---|
| cg15837522 | Chr8:117892654 | - | OpenSea | 0.556245908 | 0.514901074 | 0.036868144 | 3.10E-09 | 0.002044141 |
| cg25433259 | Chr6:159271784 | *OSTCP1* | OpenSea | 0.665418585 | 0.63835286 | 0.01226212 | 1.86E-08 | 0.003210132 |
| cg16392442 | Chr12:49595954 | - | OpenSea | 0.173533262 | 0.156960928 | 0.013945435 | 2.95E-08 | 0.003401872 |
| cg06457135 | Chr19:5953406 | *RANBP3* | OpenSea | 0.408663137 | 0.384367856 | 0.020356664 | 5.09E-08 | 0.005024759 |
| cg12910977 | Chr5:138207492 | *LRRTM2;CTNNA1* | OpenSea | 0.56139306 | 0.517909498 | 0.038321866 | 1.18E-07 | 0.007395914 |
| cg20163056 | Chr19:5005982 | *KDM4B* | OpenSea | 0.698289888 | 0.683943243 | 0.014828622 | 1.84E-07 | 0.00789425 |
| cg02023138 | Chr12:51701979 | *BIN2* | OpenSea | 0.841818365 | 0.831965078 | 0.010812848 | 2.53E-07 | 0.009729895 |
| cg20137410 | Chr22:29292645 | *ZNRF3* | OpenSea | 0.675530786 | 0.662209599 | 0.013739159 | 4.28E-07 | 0.012316665 |
| cg03629335 | Chr4:100126967 | *LOC100507053;ADH6* | OpenSea | 0.690358212 | 0.657914214 | 0.037768244 | 4.69E-07 | 0.012474048 |
| cg17687025 | Chr11:64535291 | *SF1* | S_Shore | 0.204457081 | 0.185255468 | 0.01395003 | 5.13E-07 | 0.012605011 |
| cg17745097 | Chr1:11863365 | *MTHFR* | N_Shelf | 0.083072157 | 0.076597688 | 0.008709192 | 5.36E-07 | 0.012605011 |
| cg09302321 | Chr7:141676574 | - | OpenSea | 0.423085749 | 0.401807225 | 0.019022438 | 5.85E-07 | 0.012605011 |
| cg05613718 | Chr7:38355100 | - | S_Shelf | 0.245969001 | 0.221887116 | 0.020048782 | 6.02E-07 | 0.012605011 |
| cg20911897 | Chr4:37953726 | *TBC1D1* | OpenSea | 0.468676657 | 0.452628847 | 0.02157469 | 7.96E-07 | 0.015021512 |
| cg06690548 | Chr4:139162808 | *SLC7A11* | OpenSea | 0.787899418 | 0.761387982 | 0.027485703 | 1.07E-06 | 0.017687179 |
| cg22140708 | Chr6:10081664 | - | OpenSea | 0.782067871 | 0.771703978 | 0.015494803 | 1.83E-06 | 0.022479767 |
| cg20989855 | Chr4:20985927 | *KCNIP4* | OpenSea | 0.220961686 | 0.204735145 | 0.016429303 | 1.90E-06 | 0.022643892 |
| cg03096649 | Chr7:130056977 | *CEP41* | OpenSea | 0.80181661 | 0.784614838 | 0.011533785 | 2.03E-06 | 0.022687772 |
| cg05903720 | Chr14:104663241 | - | OpenSea | 0.671042328 | 0.656044052 | 0.018513326 | 2.23E-06 | 0.022687772 |
| cg20150812 | Chr9:124029753 | *GSN* | OpenSea | 0.631561051 | 0.619989602 | 0.014187244 | 2.25E-06 | 0.022687772 |
| cg17602903 | Chr1:27986074 | - | N_Shore | 0.630259045 | 0.615597561 | 0.010167339 | 2.28E-06 | 0.022687772 |
| cg17702370 | Chr17:79283128 | *C17orf55* | N_Shore | 0.15774941 | 0.148590665 | 0.011934798 | 2.79E-06 | 0.025684613 |
| cg20218040 | Chr6:14369697 | - | OpenSea | 0.784529042 | 0.761995514 | 0.016775315 | 3.60E-06 | 0.028266416 |
| cg13039251 | Chr5:32018601 | *PDZD2* | OpenSea | 0.585971779 | 0.558399722 | 0.033387143 | 4.34E-06 | 0.030616329 |
| cg08483768 | Chr16:86304619 | - | OpenSea | 0.509458732 | 0.482372721 | 0.034480699 | 4.72E-06 | 0.032620454 |
| cg09316997 | Chr14:91874913 | *CCDC88C* | OpenSea | 0.58855649 | 0.563127383 | 0.011506387 | 5.05E-06 | 0.032907508 |
| cg03593369 | Chr2:43456557 | - | S_Shore | 0.293230936 | 0.273533246 | 0.013587607 | 5.89E-06 | 0.035030841 |
| cg21953769 | Chr19:30168190 | - | S_Shelf | 0.528806663 | 0.516187977 | 0.006921377 | 5.93E-06 | 0.035030841 |

Supplementary Table 4 (continued).

| CpG Probe | Chromosomal position[a] | Gene | Relation to Island | Cases: Mean beta value | Controls: Mean beta value | Log-fold change[b] | Raw P-value | Q-Value[c] |
|---|---|---|---|---|---|---|---|---|
| cg06548416 | Chr1:24438703 | *MYOM3* | OpenSea | 0.232424437 | 0.205557918 | 0.028630552 | 6.24E-06 | 0.036283587 |
| cg03442235 | Chr6:15369118 | *JARID2* | OpenSea | 0.721697523 | 0.709886922 | 0.016372972 | 6.75E-06 | 0.037855611 |
| cg11994115 | Chr19:40360856 | *FCGBP* | N_Shore | 0.643066668 | 0.618532712 | 0.026449583 | 6.87E-06 | 0.037855611 |
| cg15081698 | Chr6:101847050 | *GRIK2* | Island | 0.123312573 | 0.111708053 | 0.010802766 | 6.97E-06 | 0.037855611 |
| cg23852535 | Chr17:8857258 | *PIK3R5* | OpenSea | 0.837875022 | 0.83078653 | 0.007379714 | 7.32E-06 | 0.038533382 |
| cg14217303 | Chr15:85177537 | *SCAND2P* | S_Shore | 0.256459622 | 0.247047364 | 0.009832479 | 7.36E-06 | 0.038533382 |
| cg03366951 | Chr3:39302545 | - | OpenSea | 0.163064368 | 0.153998609 | 0.014080144 | 8.95E-06 | 0.042680011 |
| cg03741619 | Chr17:3438918 | *TRPV3* | Island | 0.073890023 | 0.068182814 | 0.005445805 | 1.03E-05 | 0.046105568 |
| cg01601658 | Chr6:168785524 | - | OpenSea | 0.307781447 | 0.288746997 | 0.019683281 | 1.04E-05 | 0.046456723 |
| cg08635097 | Chr13:44833857 | - | OpenSea | 0.129426117 | 0.123140906 | 0.007551291 | 1.11E-05 | 0.048688472 |
| cg21539223 | Chr5:112312093 | *DCP2* | N_Shore | 0.049278962 | 0.044646758 | 0.004760791 | 1.13E-05 | 0.048994621 |
| cg08960830 | Chr11:75047180 | *ARRB1;MIR326* | OpenSea | 0.699904812 | 0.692487485 | 0.009050025 | 1.16E-05 | 0.048994621 |

[a]Positions are based on the human reference genome assembly GRCh38.
[b]Fold change comparing beta estimates between cases and controls.
[c]False-discovery rate (FDR)-adjusted p-value.

Supplementary Table 5. CpGs used for multifactorial modeling among participants with genetic data (n=38 CpGs).

| CpG Probe | Chromosomal position[a] | Gene | Relation to Island | Cases: Mean beta value | Controls: Mean beta value | Log-fold change[b] | Raw P-value | Q-Value[c] |
|---|---|---|---|---|---|---|---|---|
| cg15837522 | Chr8:117892654 | - | OpenSea | 0.55624591 | 0.51490107 | 0.03686814 | 3.10E-09 | 0.00204414 |
| cg25433259 | Chr6:159271784 | OSTCP1 | OpenSea | 0.66541858 | 0.63835286 | 0.01226212 | 1.86E-08 | 0.00321013 |
| cg16392442 | Chr12:49595954 | - | OpenSea | 0.17353326 | 0.15696093 | 0.01394544 | 2.95E-08 | 0.00340187 |
| cg06457135 | Chr19:5953406 | RANBP3 | OpenSea | 0.40866314 | 0.38436786 | 0.02035666 | 5.09E-08 | 0.00502476 |
| cg12910977 | Chr5:138207492 | LRRTM2;CTNNA1 | OpenSea | 0.56139306 | 0.5179095 | 0.03832187 | 1.18E-07 | 0.00739591 |
| cg20163056 | Chr19:5005982 | KDM4B | OpenSea | 0.69828989 | 0.68394324 | 0.01482862 | 1.84E-07 | 0.00789425 |
| cg02023138 | Chr12:51701979 | BIN2 | OpenSea | 0.84181837 | 0.83196508 | 0.01081285 | 2.53E-07 | 0.00972989 |
| cg20137410 | Chr22:29292645 | ZNRF3 | OpenSea | 0.67553079 | 0.6622096 | 0.01373916 | 4.28E-07 | 0.01231666 |
| cg03629335 | Chr4:100126967 | LOC100507053;ADH6 | OpenSea | 0.69035821 | 0.65791421 | 0.03776824 | 4.69E-07 | 0.01247405 |
| cg05613718 | Chr7:38355100 | - | S_Shelf | 0.245969 | 0.22188712 | 0.02004878 | 6.02E-07 | 0.01260501 |
| cg09302321 | Chr7:141676574 | - | OpenSea | 0.42308575 | 0.40180723 | 0.01902244 | 5.85E-07 | 0.01260501 |
| cg17687025 | Chr11:64535291 | SF1 | S_Shore | 0.20445708 | 0.18525547 | 0.01395003 | 5.13E-07 | 0.01260501 |
| cg17745097 | Chr1:11863365 | MTHFR | N_Shelf | 0.08307216 | 0.07659769 | 0.00870919 | 5.36E-07 | 0.01260501 |
| cg06690548 | Chr4:139162808 | SLC7A11 | OpenSea | 0.78789942 | 0.76138798 | 0.0274857 | 1.07E-06 | 0.01768718 |
| cg22140708 | Chr6:10081664 | - | OpenSea | 0.78206787 | 0.77170398 | 0.0154948 | 1.83E-06 | 0.02247977 |
| cg20989855 | Chr4:20985927 | KCNIP4 | OpenSea | 0.22096169 | 0.20473515 | 0.0164293 | 1.90E-06 | 0.02264389 |
| cg03096649 | Chr7:130056977 | CEP41 | OpenSea | 0.80181661 | 0.78461484 | 0.01153378 | 2.03E-06 | 0.02268777 |
| cg05903720 | Chr14:104663241 | - | OpenSea | 0.67104233 | 0.65604405 | 0.01851333 | 2.23E-06 | 0.02268777 |
| cg17602903 | Chr1:27986074 | - | N_Shore | 0.63025904 | 0.61559756 | 0.01016734 | 2.28E-06 | 0.02268777 |
| cg20150812 | Chr9:124029753 | GSN | OpenSea | 0.63156105 | 0.6199896 | 0.01418724 | 2.25E-06 | 0.02268777 |
| cg17702370 | Chr17:79283128 | C17orf55 | N_Shore | 0.15774941 | 0.14859067 | 0.0119348 | 2.79E-06 | 0.02568461 |
| cg20218040 | Chr6:14369697 | - | OpenSea | 0.78452904 | 0.76199551 | 0.01677532 | 3.60E-06 | 0.02826642 |
| cg13039251 | Chr5:32018601 | PDZD2 | OpenSea | 0.58597178 | 0.55839972 | 0.03338714 | 4.34E-06 | 0.03061633 |
| cg08483768 | Chr16:86304619 | - | OpenSea | 0.50945873 | 0.48237272 | 0.0344807 | 4.72E-06 | 0.03262045 |
| cg09316997 | Chr14:91874913 | CCDC88C | OpenSea | 0.58855649 | 0.56312738 | 0.01150639 | 5.05E-06 | 0.03290751 |
| cg03593369 | Chr2:43456557 | - | S_Shore | 0.29323094 | 0.27353325 | 0.01358761 | 5.89E-06 | 0.03503084 |
| cg21953769 | Chr19:30168190 | - | S_Shelf | 0.52880666 | 0.51618798 | 0.00692138 | 5.93E-06 | 0.03503084 |
| cg06548416 | Chr1:24438703 | MYOM3 | OpenSea | 0.23242444 | 0.20555792 | 0.02863055 | 6.24E-06 | 0.03628359 |
| cg11994115 | Chr19:40360856 | FCGBP | N_Shore | 0.64306667 | 0.61853271 | 0.02644958 | 6.87E-06 | 0.03785561 |
| cg15081698 | Chr6:101847050 | GRIK2 | Island | 0.12331257 | 0.11170805 | 0.01080277 | 6.97E-06 | 0.03785561 |
| cg14217303 | Chr15:85177537 | SCAND2P | S_Shore | 0.25645962 | 0.24704736 | 0.00983248 | 7.36E-06 | 0.03853338 |
| cg23852535 | Chr17:8857258 | PIK3R5 | OpenSea | 0.83787502 | 0.83078653 | 0.00737971 | 7.32E-06 | 0.03853338 |
| cg03366951 | Chr3:39302545 | - | OpenSea | 0.16306437 | 0.15399861 | 0.01408014 | 8.95E-06 | 0.04268001 |

Supplementary Table 5 Continued).

| CpG Probe | Chromosomal position[a] | Gene | Relation to Island | Cases: Mean beta value | Controls: Mean beta value | Log-fold change[b] | Raw P-value | Q-Value[c] |
|---|---|---|---|---|---|---|---|---|
| cg03741619 | Chr17:3438918 | TRPV3 | Island | 0.07389002 | 0.06818281 | 0.00544581 | 1.03E-05 | 0.04610557 |
| cg01601658 | Chr6:168785524 | - | OpenSea | 0.30778145 | 0.288747 | 0.01968328 | 1.04E-05 | 0.04645672 |
| cg08635097 | Chr13:44833857 | - | OpenSea | 0.12942612 | 0.12314091 | 0.00755129 | 1.11E-05 | 0.04868847 |
| cg08960830 | Chr11:75047180 | ARRB1;MIR326 | OpenSea | 0.69990481 | 0.69248749 | 0.00905002 | 1.16E-05 | 0.04899462 |
| cg21539223 | Chr5:112312093 | DCP2 | N_Shore | 0.04927896 | 0.04464676 | 0.00476079 | 1.13E-05 | 0.04899462 |

[a]Positions are based on the human reference genome assembly GRCh38.
[b]Fold change comparing beta estimates between cases and controls.
[c]False-discovery rate (FDR)-adjusted p-value.
